# Pharmacogenomic scores in psychiatry: Systematic review of existing evidence

**DOI:** 10.1101/2024.04.05.24305376

**Authors:** Nigussie T. Sharew, Scott R. Clark, K. Oliver Schubert, Azmeraw T. Amare

## Abstract

In the past two decades, significant progress has been made in the development of polygenic scores (PGSs). One specific application of PGSs is the development and potential use of pharmacogenomic- scores (PGx-scores) to identify patients who can benefit from a specific medication or are likely to experience side effects. This systematic review comprehensively evaluates published PGx-score studies in psychiatry and provides insights into their potential clinical use and avenues for future development. A systematic literature search was conducted across PubMed, EMBASE, and Web of Science databases until 22 August 2023. This review included fifty-three primary studies, of which the majority (69.8%) were conducted using samples of European ancestry. We found that over 90% of PGx-scores in psychiatry have been developed based on psychiatric and medical diagnoses or trait variants, rather than pharmacogenomic variants. Among these PGx-scores, the polygenic score for schizophrenia (PGSSCZ) has been most extensively studied in relation to its impact on treatment outcomes (32 publications). Twenty (62.5%) of these studies suggest that individuals with higher PGSSCZ have negative outcomes from psychotropic treatment: poorer treatment response, higher rates of treatment resistance, more antipsychotic-induced side effects, or more psychiatric hospitalizations, while the remaining studies didn’t find significant associations. Although PGx-scores alone accounted for at best 5.6% of the variance in treatment outcomes (in schizophrenia treatment resistance), together with clinical variables they explained up to 13.7% (in bipolar lithium response), suggesting that clinical translation might be achieved by including PGx-scores in multivariable models. In conclusion, our literature review found that there are still very few studies developing PGx-scores using pharmacogenomic variants. Research with larger and diverse populations is required to develop clinically relevant PGx-scores, using biology-informed and multi-phenotypic polygenic scoring approaches, as well as by integrating clinical variables with these scores to facilitate their translation to psychiatric practice.

## Introduction

Psychiatric disorders are significant contributors to the global disease burden and represent a major public health concern ^1^, highlighting the urgent need for effective prevention and treatment strategies ^2^. The 2022 World Health Organization (WHO) report estimates that nearly a billion people suffer from psychiatric disorders, with an associated economic loss of $2 trillion per year and this figure is expected to rise to $6 trillion by 2030 ^3–6^.

Pharmacological treatments including antidepressants, antipsychotics, mood stabilizers, and anxiolytics are commonly prescribed for people suffering from psychiatric disorders ^7^.

However, the effectiveness of these medications varies between individuals, with some responding well while others do not show notable improvement or experience adverse effects^7^. For example, among patients with major depressive disorder (MDD), 30-40% fail to respond to the first-line pharmacological treatment options of selective serotonin reuptake inhibitors (SSRIs), and 10-45% exhibit moderate to severe treatment-related side effects ^8, 9^.

Similarly, only 30% of patients with bipolar disorder (BD) show a full clinical response to first-line lithium monotherapy ^10^, and up to 25% of patients with first-episode schizophrenia (SCZ) are treatment-resistant to first-line antipsychotics ^11^. This variability in pharmacological treatment outcomes can be attributed to the complex interplay of genetic and environmental factors, including patients’ clinical characteristics (e.g., severity, number, and duration of illness episodes), as well as sociodemographic variables ^12^. For example, in individuals with MDD, genetic factors account for 42-52% of the observed differences in antidepressant treatment response, while environmental factors contribute to the remainder ^13,14^.

To date, studies employing both candidate gene investigations (pharmaco*genetics*) and genome-wide (pharmaco*genomic*) approaches, have successfully pinpointed genetic variations associated with treatment outcomes in psychiatry, including response ^15^, remission ^16^, resistance ^17^ and adverse drug reactions ^18^. For instance, the pharmaco*genetic* approach has uncovered genetic polymorphisms within genes encoding drug-metabolizing enzymes including those involved in the metabolism of various psychotropic drugs (e.g., *CYP2D6* and *CYP2C19*) ^19^ as well as drug transporters (e.g., *5-HTTLPR),* establishing their association with patients’ responses to medications^20^. This evidence is now incorporated into commercially available pharmacogenetic testing panels, aiding drug selection and dose adjustments and ultimately aiming at improving medication efficacy and tolerability^21, 22^.

Similarly, the pharmaco*genomics* approach has revealed a number of genetic polymorphisms located within or near pharmacologically relevant candidate genes that influence individuals’ reaction to psychiatric medications ^10^. For instance, Hou et al identified four linked genetic variants on chromosome 21 associated with lithium response in a Genome-wide Association Study (GWAS) ^10^. It has been challenging, however, to translate these pharmaco*genomic* findings into clinical practice, mainly due to the small effect size of individual genetic variants on treatment outcomes, along with a limited understanding of gene function ^23^.

In an effort to improve effect estimates and make pharmaco*genomic* findings more clinically relevant, researchers have recently adopted polygenic score methods combine the effect of multiple genetic variants across the genome and have developed pharmacogenomic scores (PGx-scores) ^24, 25^. In this systematic review, we provide a detailed account of the research undertaken to date, and of the performance, shortfalls, and future recommendations for the development of PGx-scores for the personalisation of psychiatric care.

## Methods

This systematic review adhered to the PRISMA updated guidelines 2020 ^26^ and was registered with the International Prospective Register of Systematic Reviews (PROSPERO) on February 9, 2023 (ID = <u>CRD42023395404</u>). The review protocol was prepared before commencement to ensure a transparent and standardized methodology.

### Search strategy, inclusion, and exclusion criteria

The literature search was performed across three databases including PubMed, EMBASE, and Web of Science databases from January 1^st^, 2005 to 22^nd^ August 2023, by using search string: ((“Polygenic score*” OR “Polygenic risk score*” OR “Risk profile score*” OR “Genetic risk score*” OR “Gene score*” OR “Genetic score*” OR polygenic* OR “Pharmacogenomic variants” OR “Pharmacogenomic testing” OR Pharmaco-omic* OR pharmacogeno* OR “Pharmacogenetics”) AND (“Antipsychotic agents” OR antipsycho* OR “Antidepressive agents” Antidepress* OR “Anti-anxiety agents” OR Anti-anxiet* OR Valproic acid OR Valproate OR Divalproate OR Divalproex OR Carbamazepine OR Oxcarbazepine OR Risperidone OR Gabapentin OR Lamotrigine OR Licarbazepine OR Pregabalin OR Tiagabine OR Zonisamide OR Lithium)) Our search strategy included all original studies that developed PGx-score for drug-related phenotypes such as, drug dosage, therapeutic drug response, resistance, drug-induced side-effects, relapse or hospitalisation in psychiatry. We included studies that reported weighted PGx-score for the drug-related phenotypes mentioned above, while excluding publications in languages other than English, conference abstracts, case reports, editorials, notes, and systematic reviews. NTS screened the studies for inclusion under supervision of ATA. In the final step, all studies were imported into Endnote version 20, a reference manager software. Duplicate entries were removed, and the selection of studies was carried out based on the predetermined inclusion and exclusion criteria. Supplementary file 1 provides details of the systematic search strategies and results in each database.

*Insert Supplementary file 1 here*.

### Data extraction and synthesis

NTS extracted data using a customised data extraction excel sheet format, under supervision of ATA. This excel sheet included information on the authors’ characteristics, details of the drug outcomes, characteristics of the study cohort (such as base, target, and validation cohorts), number of variants included in the polygenic score (PGS), polygenic scoring methods, and association effect estimates. The “target cohorts” describe the cohorts where the PGS was developed and tested, while “discovery cohorts” refers to the cohorts utilized to create GWAS summary statistics. “Validation cohorts” are independent cohorts where the PGSs were validated. “Variance explained” measures the proportion of phenotype variance the PGS can account for in a predictive model assuming linear effects. Coefficient of effect estimates, standard error, and sample size were used to calculate odd ratios if not reported in the studies. The results were organized thematically based on the psychiatric disorders that were studied, as well as the specific phenotypes investigated, including treatment response, treatment resistance, and drug-induced side effects.

*Insert Supplementary file 2 here*.

### Quality assessment

The quality of included studies was assessed using a quality assessment form adapted from previously validated and published sources ^27, 28^. The assessment criteria covered various aspects of the study design, such as the rationale and methods of PGS, power calculation, inclusion and exclusion criteria, basic characteristics of the study population, availability of validation cohort, type of analysis, correction for multiple testing, and consideration of confounders in the analysis. The quality assessment was conducted by NTS under supervision of ATA.

## Results

Our initial search identified a total of 4,889 studies that were potentially relevant to the research topic. After removing 1,586 duplicated publications, 3,303 articles remained for the title and abstract screening. Subsequently, 3,175 studies were excluded during the initial title and abstract screening phase, leaving 127 articles for full text review. Finally, 53 studies met the predetermined inclusion criteria and were included in the final synthesis. Figure 1 presents the flowchart of the step-by-step process of study selection with reasons for exclusion.

**Figure 1.**
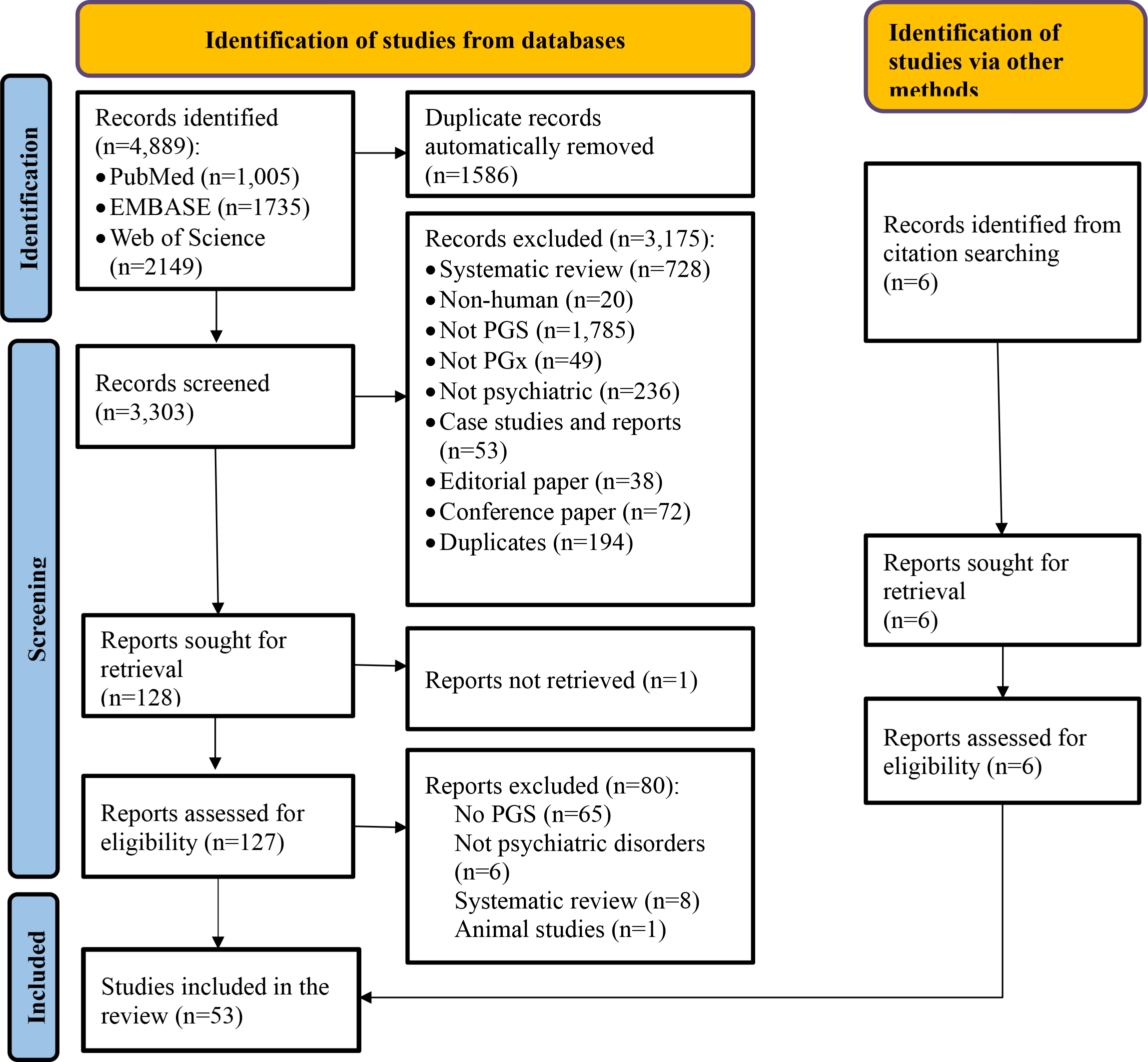
PRISMA flow diagram showing the steps of screening studies included in this systematic review. *Abbreviations:* PGS = Polygenic score; PGx =Pharmacogenomics

### Quality assessment

Nearly three-quarters (39/53) of studies described the rationale for the selected polygenic scoring methods, while about 20% (10/53) of studies performed a power calculation. All studies reported the inclusion and exclusion criteria for participants’ selection. Only fourteen studies used external cohorts to validate their findings. Correction for multiple testing was performed in 83.2% (44/53) of studies. Detailed results of the quality assessment are provided in a supplementary file 1.

Most studies 37 (69.8%) were conducted on samples comprising individuals of European ancestry. Eleven studies (20.8%) included participants from other ancestries, such as African, African American and/or East Asian. Three studies targeted only Latin American participants and another two studies were conducted specifically on samples of East Asian ancestry.

However, there was no study solely centered on samples of African ancestry. A combined analysis of both the target and discovery samples showed that 14,893,321 (90%) of participants had European descent, with an increased trend over the years 2013-2023, both in the target (Figure 2A) and discovery cohorts (Figure 2B).

**Figure 2.**
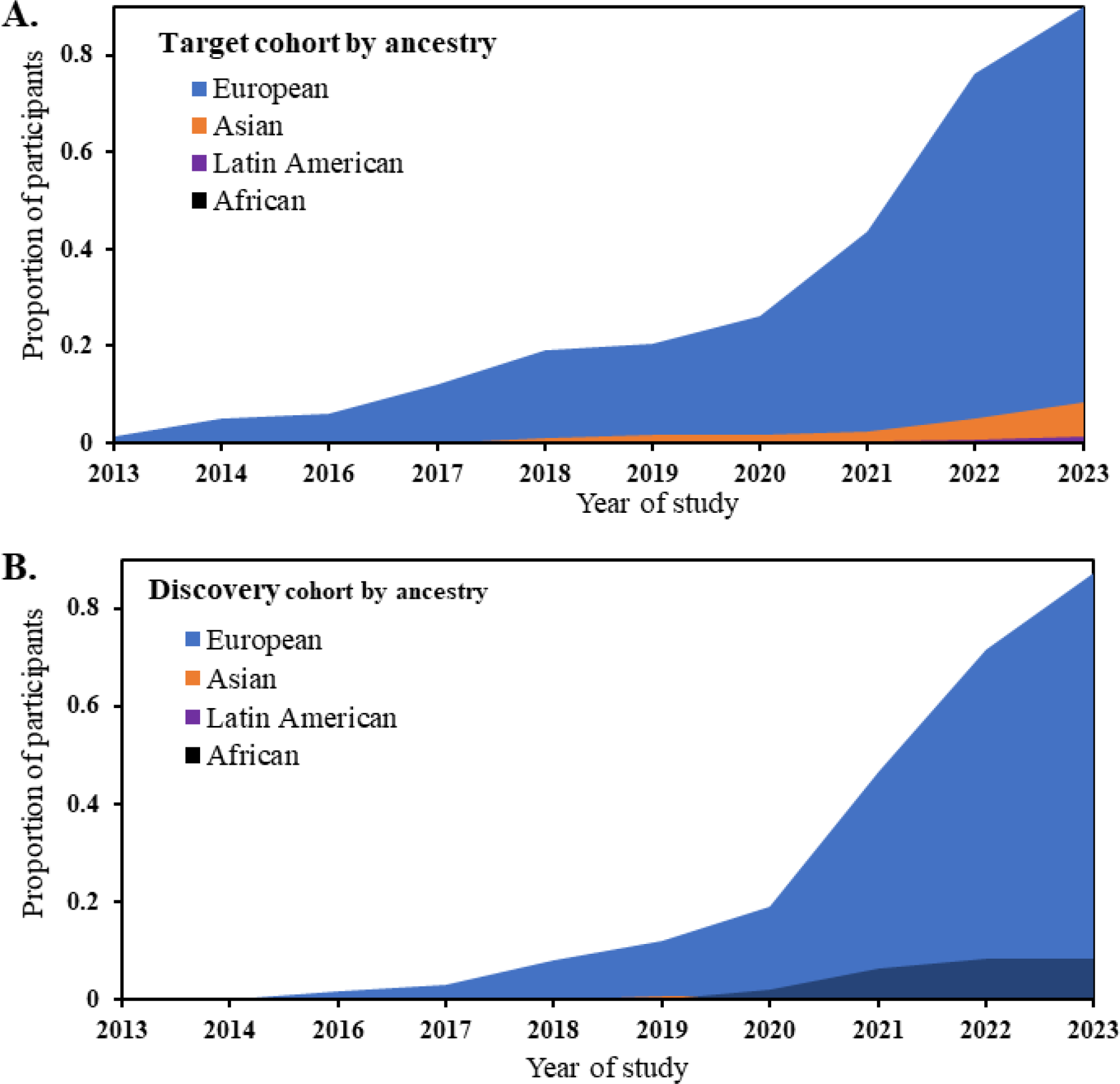
The ancestry characteristics of samples in the A) target and B) discovery cohort for studies from 2013 – 2023

The sample sizes across the studies varied widely, ranging from 44 participants ^29^ to 12,863 participants ^30^ with a median sample size of 863 in the target cohorts. Three major psychiatric conditions namely SCZ, depression and BD were the focus of included studies. In the case of SCZ, nearly 80% (21/27) of studies investigated the association between PGS and response to second-generation antipsychotics (clozapine, risperidone, lurasidone, olanzapine, aripiprazole, quetiapine, ziprasidone, and perphenazine). About half of SCZ studies (13/27) exclusively analysed clozapine treatment outcome. Nearly three-quarters of the included studies involving patients MDD (14/19) considered the relationship between PGS and SSRIs such as citalopram or escitalopram. Six out of seven included studies developed PGx-scores and examined their associations with lithium treatment response in patients with BD.

### The association of pharmacogenomic scores with treatment outcomes

Involving patients with psychiatric disorders, researchers have developed several PGx-scores and investigated their associations with key treatment outcomes: treatment response, treatment resistance, treatment-related side effects, and hospitalization rates. Table 1 provides a summary of the findings extracted from each of the articles included in this systematic review (Table 1).

**Table 1.**
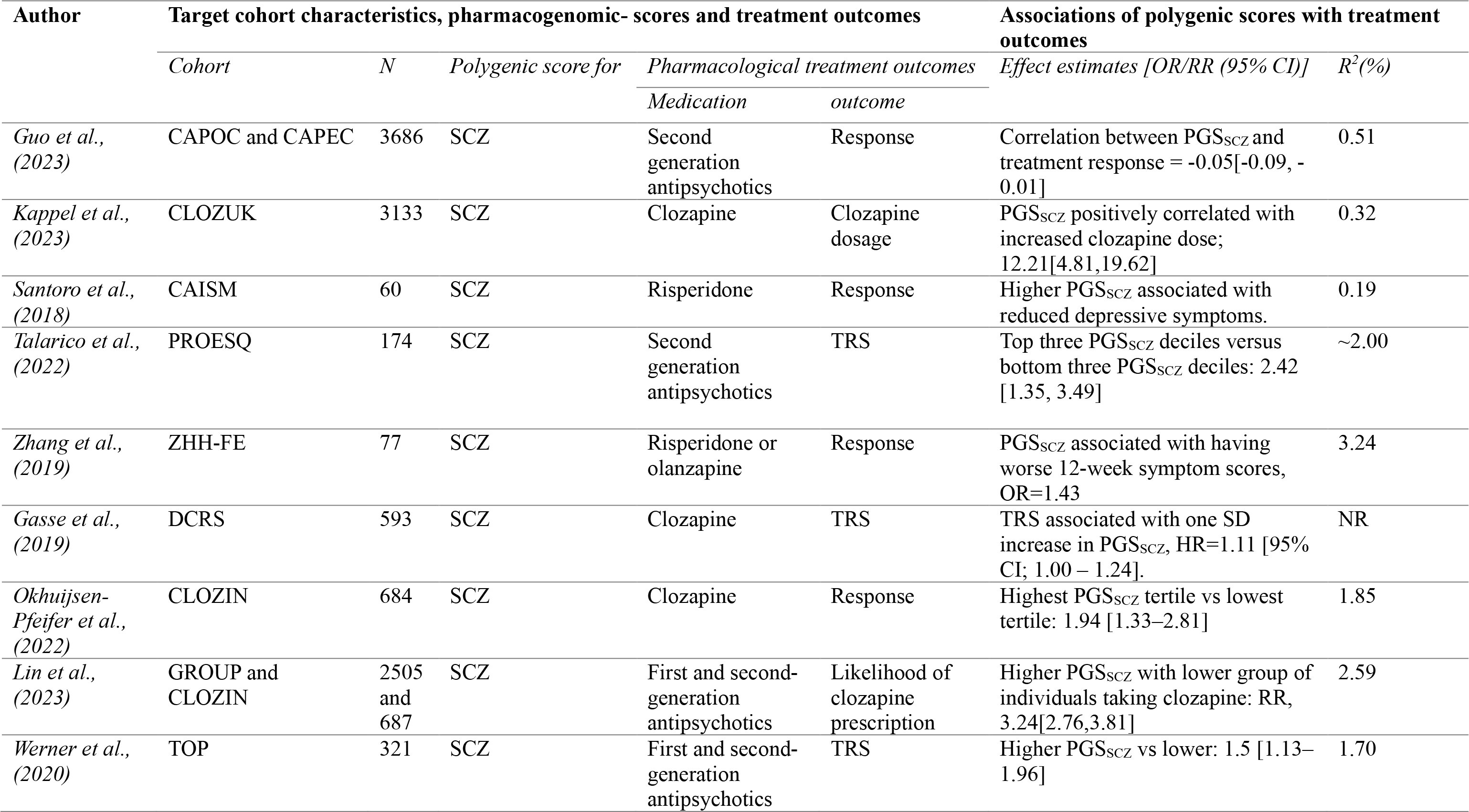
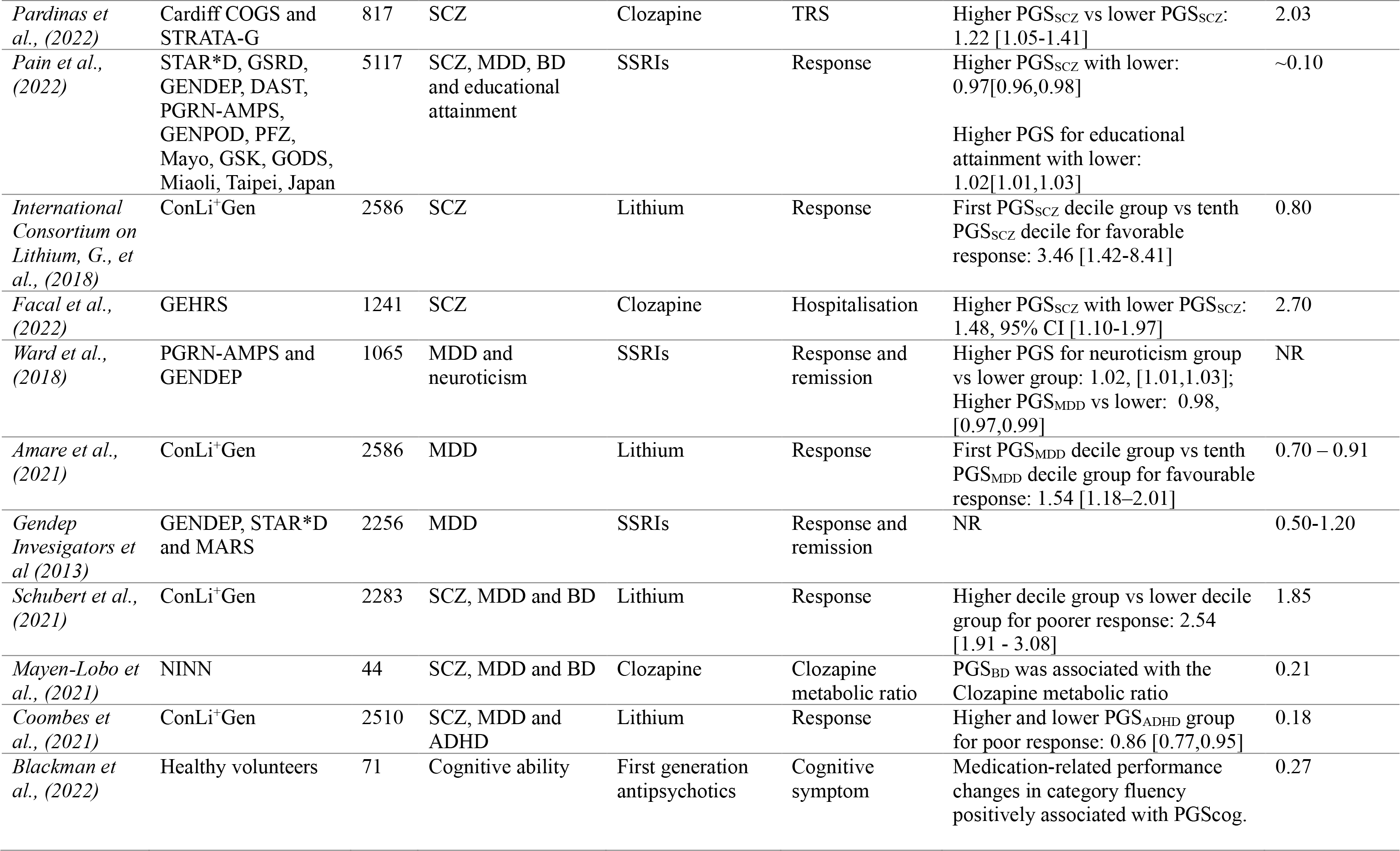
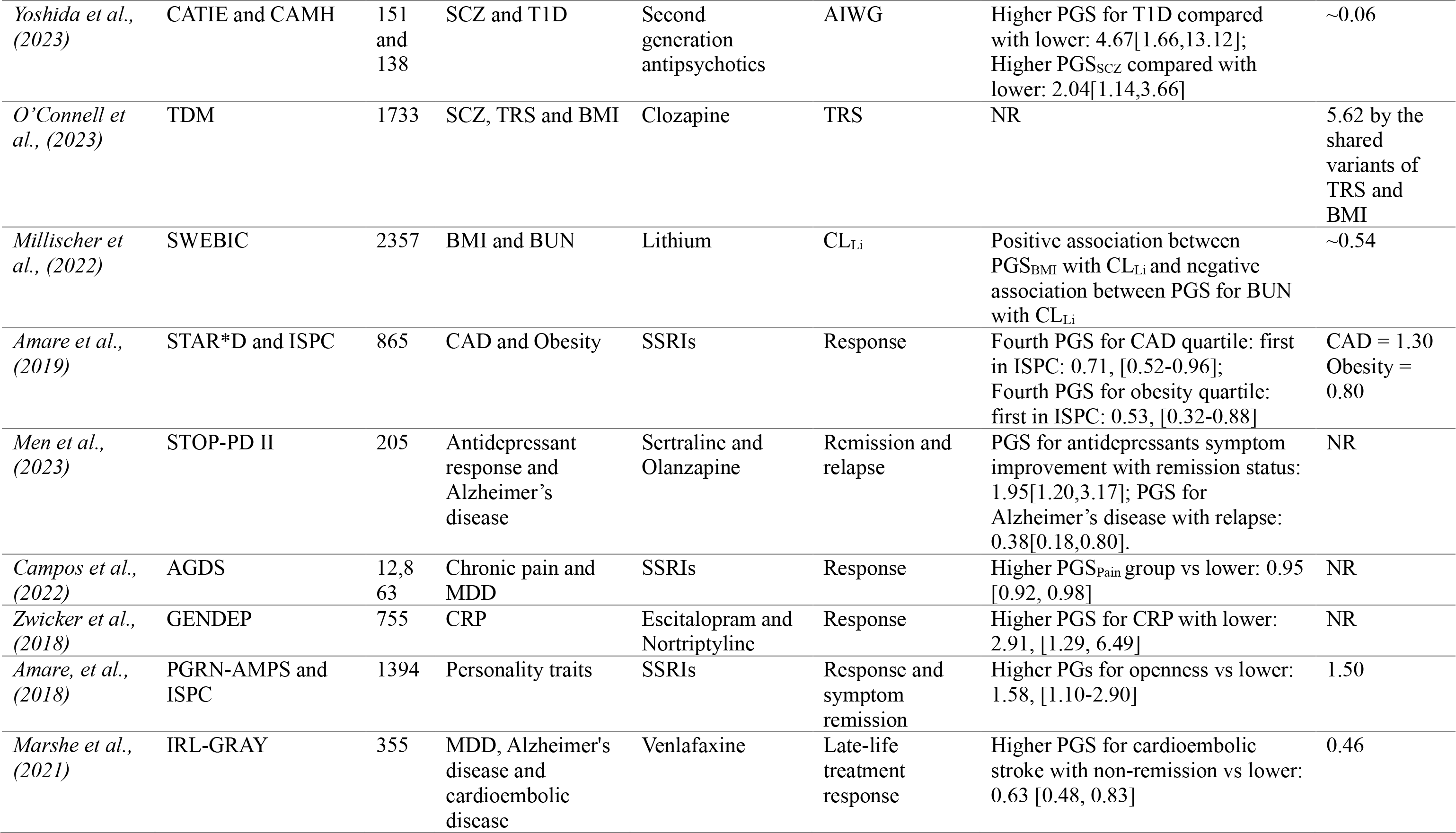
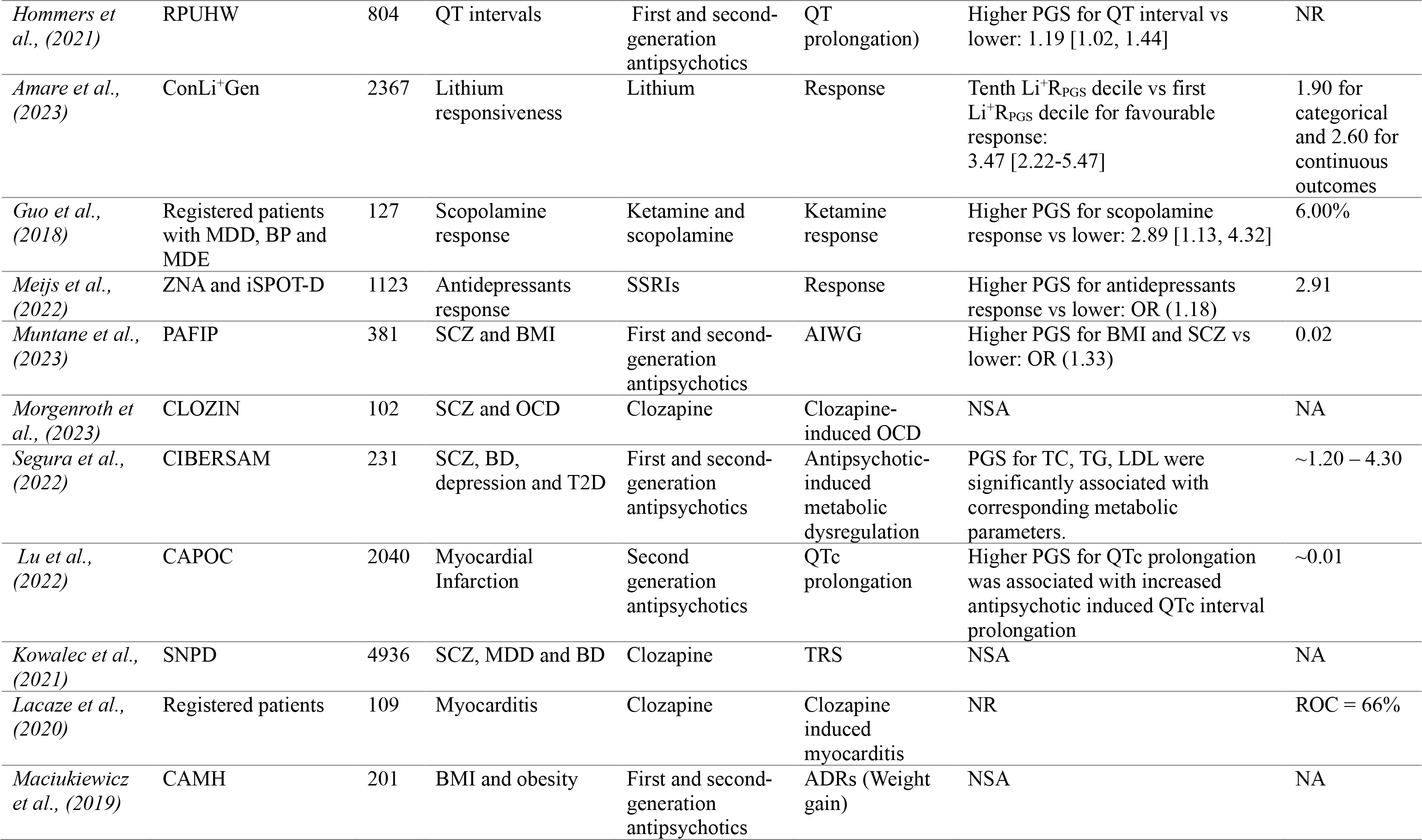
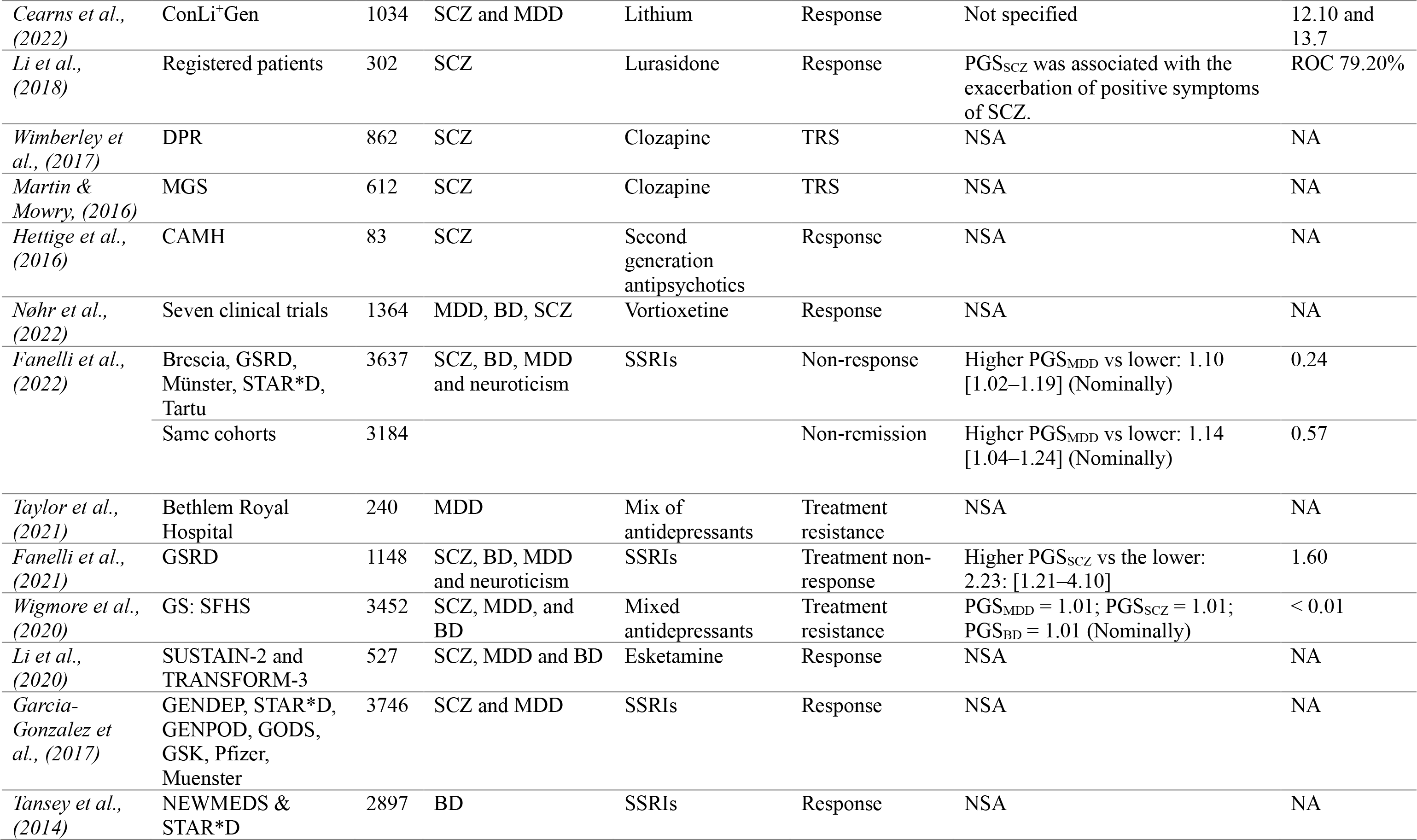
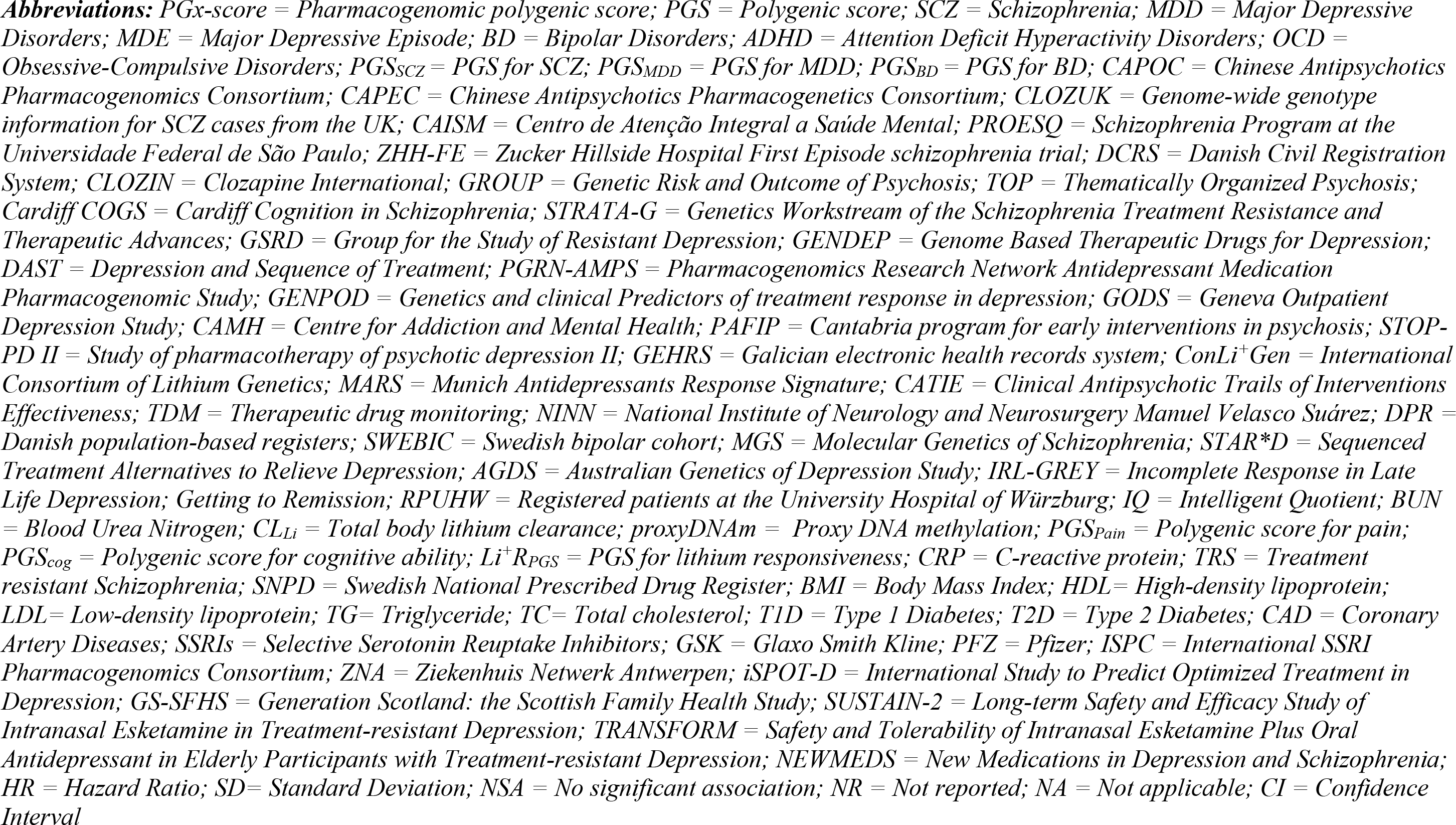
Summary of findings on the association between PGx-score and treatment outcomes.

Of all PGx-scores, the polygenic loading for schizophrenia (PGSSCZ) has been most extensively studied (32 publications) in relation to its influence on treatment outcomes. Several studies (20 studies) revealed that individuals with higher genetic loading for SCZ had a poorer treatment outcome to psychiatric medications ^31–50^, while the remaining studies didn’t find significant associations. For example, in patients with SCZ, a negative correlation (*r* = -0.05[95%CI: -0.09 – -0.01]) was found between PGSSCZ and response to second generation antipsychotics (olanzapine, aripiprazole, risperidone, quetiapine, haloperidol, ziprasidone, perphenazine) following six weeks treatment ^34^. Kappel et al.(2023) observed a positive correlation (*β*=12.21; 95%CI: 4.81 – 19.62) between PGSSCZ and high clozapine dosing (>600 mg/day) suggesting that individuals with a higher PGSSCZ may require increased doses of clozapine to achieve effective treatment response ^33^. In patients treated with risperidone, those who had a higher PGSSCZ reported more depressive symptoms ^45^, and worsened positive and negative psychotic symptoms ^46^. A higher PGSSCZ was associated with a poor response to olanzapine or risperidone OR=1.43[95%CI:1.19 – 1.67] ^43^ and an increase of one standard deviation in PGSSCZ was associated with an approximately 11% increase in the risk of developing treatment-resistant schizophrenia (TRS) (OR = 1.11[95%CI: 1.00 – 1.24]) ^44^. Patients with a higher polygenic load for SCZ were 1.22 times [95%CI: 1.05 – 1.41; R^2^ = 2.03%] more likely to be resistant to clozapine ^36^, had 1.50 times [95%CI: 1.13 – 1.96; R^2^ = 1.70%] higher odds to experience resistance to other antipsychotics ^42^ and were more likely to develop antipsychotic-induced weight gain (AIWG) ^48, 49^.

In decile-based comparisons, patients in the top three PGSSCZ deciles had a 2.42 [95%CI: 1.35 – 3.49; R^2^ ∼2.00%] times higher odds of poor response to various antipsychotic medications (olanzapine, risperidone, quetiapine, and clozapine) ^35^ and the odds of treatment resistance for those in the 8^th^ PGSSCZ decile was 6.50 times [95%CI: 1.47 – 28.80] higher than for patients in the 1^st^ decile ^35^. Patients with a higher PGSSCZ had 1.48 times [95%CI: 1.10 - 1.97; R^2^ = 2.70%] higher odds of psychiatric hospitalizations and were hospitalized longer ^39^.

Interestingly, in a study by Okhuijsen-Pfeifer et al (2022), patients treated with clozapine who were in the highest PGSSCZ tertile group were 1.94-fold more likely to experience low (i.e., more favourable) symptom severity [95%CI: 1.33 – 2.81; R^2^ = 1.85%], compared to those in the lowest PGSSCZ tertile group ^38^.

Similar to the above findings, studies involving patients with MDD and BD have also unveiled the association of a higher PGSSCZ with poorer response to either antidepressants in MDD (OR = 0.97 [95%CI: 0.96 - 0.98; R^2^ ∼0.01%]) ^37^ or lithium treatment in BD (OR = 0.29 [95%CI: 0.12 – 0.70; R^2^ = 0.80]) ^47^.

Aside from the PGSSCZ, several other polygenic scores have also been investigated for their potential to predict treatment outcomes in patients with psychiatric disorders. These include polygenic scores for cognitive function, cardiometabolic traits, MDD, BD, ADHD, autism spectrum disorders (ASD), anxiety, alcohol use disorders, personality traits, and educational attainment. In patients with SCZ, a higher genetic loading for general cognitive ability was associated with better cognitive function following antipsychotic treatment ^51^. SCZ patients carrying a greater genetic load for higher body mass index (BMI) were at a higher risk of being resistant to clozapine treatment ^31^. Moreover, the polygenic loading for BD (PGSBD) was found to be significantly associated with clozapine metabolic ratio ^29^, a measure of how clozapine is metabolized within the body, which may impact treatment response or adverse effects. In patients with first episode psychosis, higher genetic loadings for HDL, LDL, and total cholesterol predicted antipsychotic-induced metabolic disturbance ^52^.

Studies of PGx-scores for patients with MDD also explored the impact of the polygenic scores for personality traits and physical illnesses on treatment outcomes. In a study by the Genome-Based Therapeutic Drugs for Depression (GENDEP) investigators, the polygenic loading for MDD (PGSMDD) was significantly associated with response and remission to SSRIs and TCAs treatment in patients with MDD, although the direction of association was not reported ^53^. A study that assessed the relationship between PGS for various personality traits and response to SSRIs (citalopram, escitalopram, fluvoxamine) ^54^ found that a higher genetic loading for openness personality trait was associated with a better SSRIs treatment response after 8-week of treatment OR=1.58 [95%CI, 1.10-2.90] while the PGS for neuroticism was negatively associated with SSRIs treatment response ^54^. Genetic loading for cardiometabolic disease has also shown associations with response to antidepressant treatment: Marshe et al (2021) used a PGS for cardioembolic stroke to predict response to venlafaxine, an antidepressant of the serotonin-norepinephrine reuptake inhibitors (SNRI) class, after 12-week treatment. They found that a one standard deviation increase in PGS for cardioembolic stroke was associated with a decreased probability of remission (OR = 0.63[95%CI:0.48 to 0.83]) or worsened disease symptoms (Montgomery-Asberg Depression Rating Scale (MADRS), β = -5.51[95%CI: -9.45 to -1.57]) ^55^. In a different study, individuals with the highest PGSs for coronary artery disease (4^th^ quartile) had 0.53 times ([95%CI, 0.35 – 0.81]) less likelihood of experiencing favourable response to SSRIs (citalopram, escitalopram, fluvoxamine) compared to those in the 1^st^ quartile ^56^. Similarly, those with higher genetic loading for obesity (4^th^ quartile) had a 0.53 times ([95%CI, 0.32 – 0.88]) lower likelihood of achieving a positive response to SSRIs treatment ^56^. In individuals with MDD (n=5218) treated with SSRIs (citalopram, escitalopram) or a TCA (nortriptyline), the PGS for educational attainment was positively associated with SSRI response ^37^. In a cohort of patients with psychotic depression treated with sertraline and olanzapine for 36 weeks, those who had a higher polygenic loading for Alzheimer’s disease had a decreased likelihood of relapse (OR = 0.38; [95%CI: 0.18 – 0.80]) during the study period ^57^. Higher PGS for chronic pain was negatively associated with treatment response to SSRIs, tetracyclic antidepressants (mirtazapine), and SNRIs (desvenlafaxine) (OR= 0.95[95%CI: [0.92 – 0.98]) ^30^, while a higher PGS for C-reactive protein was associated with a better response to escitalopram (OR = 2.92[95%CI: 1.30 – 6.49]), but worse response to nortriptyline ^58^.

In patients with BD, those with a low PGSMDD (first decile) were 1.54 times [95%CI: 1.18 – 2.01; R^2^ = 0.91%] more likely to respond favourably to lithium than those who had high depression genetic loading (10^th^ decile) ^59^. Similarly, a higher PGS for ADHD was associated with unfavourable lithium response (OR = 0.86[95%CI: 0.77 - 0.95], R^2^ = 0.18) ^60^. Further studies using the same dataset have shown that a combined analysis of the PGSs of multiple phenotypes and PGS with patients’ clinical data can improve the predictive capacity of polygenic models. For example, a meta-analysis of the association results of the PGSSCZ and PGSMDD provided improved response prediction compared to single disorder PGS ^41^. By applying machine learning methods, the PGSSCZ and PGSMDD were combined with clinical data which resulted in an explained variance of 13.7% in lithium treatment response ^40^. In a recent study, lithium clearance, an essential parameter for maintaining therapeutic levels of lithium and adjusting dosage, was positively associated with the PGSs for BMI and estimated glomerular filtration rate (eGFR), while it was negatively associated with the PGSs blood urea nitrogen (BUN) ^61^.

In contrast to the above studies in which PGx-scores were developed based on diseases or related phenotype variants, a few recent studies used pharmacogenomic variants to calculate PGx-scores, directly indexing treatment outcome phenotypes. For instance, in cohort of patients with TRS, a PGx-score for *clozapine resistance* predicted 4.96% of the rate of TRS variation ^31^. In patients with psychotic depression treated with sertraline and olanzapine, those with a higher genetic loading for *antidepressant remission and response* had 1.95 times [95%CI: 1.20 – 3.17] higher odds of reaching remission after 36 weeks ^57^. In similar context, PGS for *response to SSRIs* (escitalopram, sertraline, venlafaxine) predicted antidepressant treatment response in patients with MDD ^62^. A study by Guo et al. (2018) utilized variants ranked by their strength of association with ketamine response, a glutamate-modulating antidepressant used in patients with Treatment-Resistant Depression (TRD), to predict scopolamine treatment response in patients with either MDD or BD who had a current major depressive episode ^63^. Findings indicated that patients with higher genetic loadings for *ketamine response* had better responses to scopolamine, an emerging antidepressant with effects on acetylcholine (Ach) neurotransmission ^63^. A polygenic score developed for *lithium treatment response* (Li^+^RPGS) within ConLi^+^Gen was evaluated in a hold-out subsample and a smaller independent replication cohort. This analysis revealed that individuals in the highest Li^+^RPGS decile were 3.47 times [95%CI: 2.22 – 5.47, R^2^ = 2.60] more responsive to lithium compared to those in the lowest PGS decile, and a linear relationship was observed across the various deciles ^64^. Figure 3 summarized the relationship between different PGx-scores of different traits and pharmacotherapeutic outcomes.

**Figure 3.**
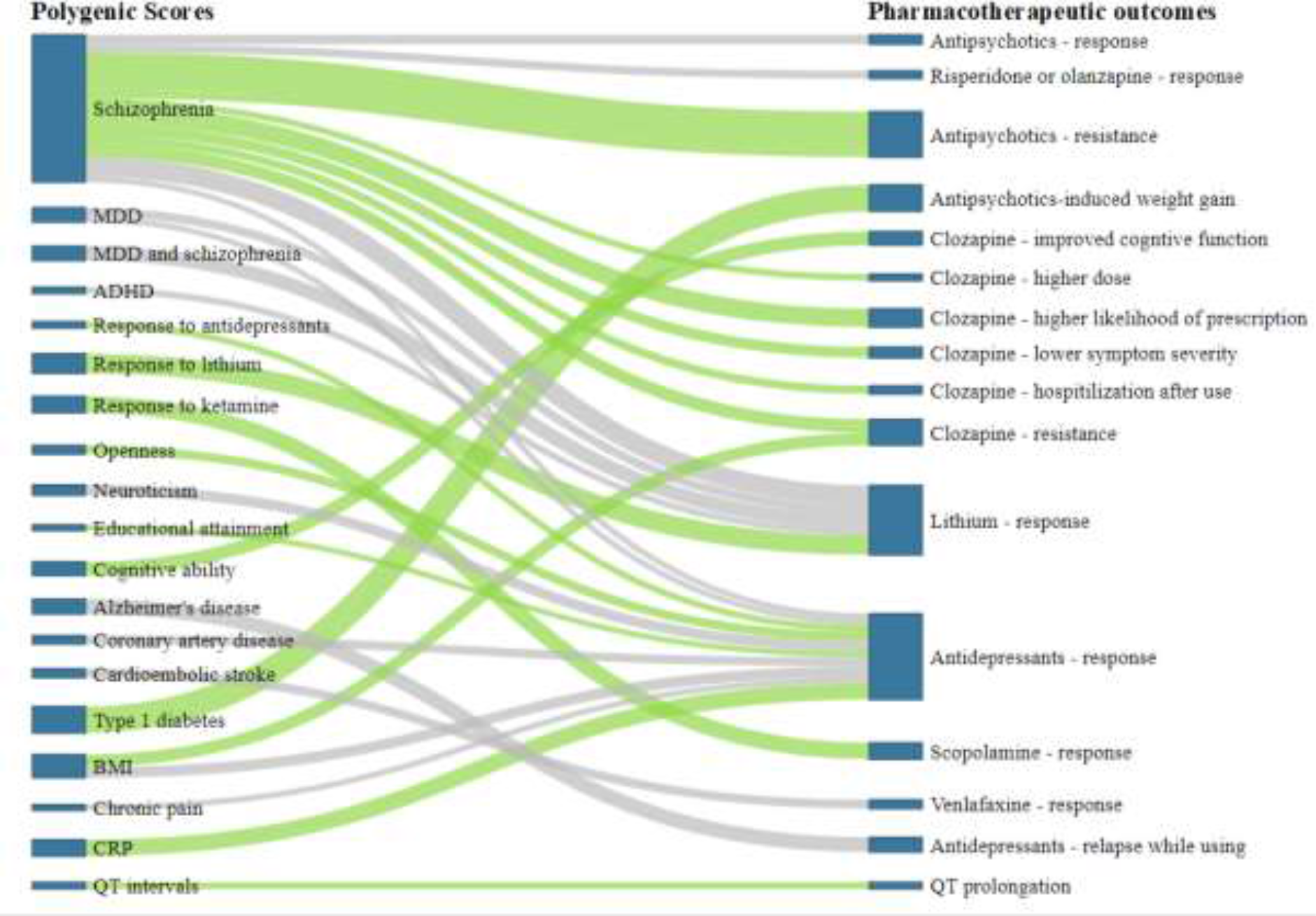
Figure showing the relationship between different PGx-scores of different traits and pharmacotherapeutic outcomes in psychiatry. *Legend:* Green line represents the positive associations of PGx-scores with treatment outcomes; Gray line indicates negative associations between PGx-scores and treatment outcomes. A wider (thick) line lines represent a stronger association.

## Discussion

Pharmacogenomic scores (PGx-scores) are emerging as novel tools for predicting treatment outcomes in psychiatry such as response, remission, resistance, side effects, or hospitalization rates. While the bench-to-bedside translation of PGx-scores has not yet been achieved, a growing body of evidence indicates their potential clinical use for treatment personalisation. In this systematic review, we describe the landscape of 53 PGx-score studies in clinical psychiatry. These PGx-scores have been developed either from genetic variants associated with psychiatric or medical diagnoses (the majority of studies); or from pharmacogenomic variants associated with treatment outcome phenotypes (a few recent studies). Individual PGx-scores alone do not explain enough variance in clinically relevant outcomes and their combination with clinical data and/or other biological markers is required for effective translation.

First, we found that over 90% of PGx-scores have been developed based on genetic variants of psychiatric or medical diagnoses (e.g., SCZ, MDD, BD, ADHD, coronary artery disease (CAD)) or phenotypes related to diagnoses (e.g., cognitive function, personality traits, educational attainment, CRP level, BMI). Among these, the PGSSCZ has been most extensively studied and has consistently shown association with pharmacotherapeutic outcomes across drug classes including antipsychotics, antidepressants and lithium, explaining as much as 3.2% of interindividual variability in some treatment outcomes ^43^. The consistent association of the PGSSCZ and treatment outcomes may be attributed to two factors. First, SCZ has a strong genetic basis with a heritability estimate of 80-85% ^65^ and it is possible that PGSSCZ captures a substantial amount of the phenotypic variance of the disorder. Previous studies have shown a direct correlation between a higher phenotypic heritability and a better predictive power of PGS ^66^. Second, SCZ GWASs are well powered, including cases and controls of diverse ancestral background ^67, 68^, leading to more accurate PGSs ^69^. The size of GWAS discovery samples has been associated with a better accuracy and predictive power of PGSs ^69^. For example, the Psychiatric Genomics Consortium (PGC in 2009) found that common genetic variants explained only 3% of the total variance in risk to SCZ, in a sample of 3322 individuals with SCZ and 3587 controls of European ancestry^70^. In a follow up study (in 2014) with expanded sample size and diversity (36,989 cases, 113,075 controls, multiple cohorts of East Asian ancestral background), the variance explained by PGSSCZ substantially increased to around 18% ^71, 72^.

It is important to highlight that in most of the reviewed studies, high PGSSCZ was associated with *poor* treatment response ^32–34, 37, 38, 40, 41, 43, 45–47^, more treatment resistance ^31, 35, 36, 42, 44^, more antipsychotics-induced side effects ^48–50^ or more psychiatric hospitalizations ^39^. A notable exception was a positive association with lower symptom burden in SCZ patient treated with clozapine ^38^. A possible explanation is that a high PGSSCZ loadings may index individuals with a higher neurodevelopmental contribution to mental disorder aetiology. Neurodevelopmental hypotheses are well established in SCZ, for instance the excessive synaptic pruning linked to complement system genotype ^73^. Psychosis prodrome and onset ^74, 75^ and TRS ^76^ has been linked to reduced brain volume and connectivity. These ‘hard wired’ brain characteristics may be more difficult to influence therapeutically through first-line (e.g., non-clozapine) pharmacological strategies ^76^.

The review also identified polygenic associations between cardiometabolic disorders ^55, 56^, personality traits ^54^, and treatment outcomes. A higher PGSs for CAD, obesity, and neurotic personality were associated with poor response to antidepressants ^54, 56^ while a positive association was found with the PGS for openness personality ^54^. This is possibly due to shared biological mechanisms, for example, a genetic overlap between major psychiatric disorders and cardiometabolic diseases ^77–80^, neuroticism ^81^, or openness personality traits ^82^ and also associated multimorbidity across these disorders ^83^ that might impact patients treatment outcomes. Personality traits have an impact on medication adherence, with neuroticism linked to non-adherence and openness to compliance ^84^. These findings indicated that disease related PGS may help us to understand underlying pathology and identify drug targets, however, there may be limitations in their utility for pharmacogenomic testing due to challenges of interpretation.

Second, from our review, it is clear that there is a major research gap regarding PGx-scores developed from pharmacogenomic variants ^31, 57, 62–64^. The lack of these studies is associated with the limited availability of well powered GWAS summary statistics on treatment outcomes (target sample) and challenges to collect genetic and clinical data from patients of a specific diagnoses, treated with similar medications (discovery sample). Currently, large scale GWASs leverage biobank datasets, where there is limited phenotyping on medication, missing standardised data on treatment outcomes.

Although the current cohort sizes for PGx-score development are much smaller than those of large scale diagnosis-based GWASs, promising initiatives are underway to achieve deeper phenotyping for medications such as lithium ^85^, clozapine ^36, 86^, and antidepressants ^37^. For instance, ConLi^+^Gen cohort, which aimed to study the genetics of lithium treatment response in individuals with BD, currently has a sample size of 2367 patients of European ancestry and 220 patients of Asian ancestry with current effort underway for a larger more diverse cohort and more detailed phenotyping ^85^. By expanding current efforts, there may be opportunities to develop pharmacogenomic PGx-scores with improved accuracy for clinical use.

The third finding from this review is that PGx-score alone fall short of explaining adequate variance in treatment outcomes for clinical translation. Notably, the highest reported explained variance solely attributed to PGx-score, by leveraging genetic variants of TRS and BMI, was 5.6% in resistance to clozapine. To address this shortfall, the combination of PGx-scores with clinical data could potentially enhance clinical use. For instance, a study modelled PGSSCZ + PGSMDD with patients’ clinical characteristics using machine learning, was able to explain 13.7% of the variance in lithium treatment responses ^40^. A further example is a multimodal model combining PGS with socio-demographic, clinical, biomarkers and structural imaging to predict rehospitalization risk showed a negative predictive value of 81.57% compared with a PGS-only model (54.83%) ^87^. Similarly, a study that modelled polygenic scores of SCZ, MDD, and BD, along with proxy DNA methylation data and clinical symptom variables showed good regression performance for prediction of response to multiple antipsychotic drugs (ROC = 0.87 [95% CI: 0.87-0.88] ^34^. In patients with type 2 diabetes, combining PGS with clinical data such as smoking status, BMI, blood lipid levels, blood pressure, and the use of anti-hypertensive and lipid-lowering medications, substantially improved the accuracy in classifying individuals into low-, moderate-, and high-risk categories for cardiovascular events to 83%, whereas accuracy was 58% with PGSs alone (29 optimized univariable PGS) ^88^. It is evident from these studies that PGx-score can be clinically useful if prediction models are refined based on a combination of PGx-scores and clinical data.

## Limitations

Some of the limitations of the present systematic review should be highlighted. First, the study participants of the studies were predominantly drawn from European populations that limits the ability to apply study’s conclusion to non-European populations and raising concerns about the generalizability of the findings to more diverse populations. Second, the inconsistent reporting of the polygenic model parameters across studies makes it challenging to compare the efficacy and obscure the true picture of PGx-score in predicting psychopharmacotherapeutic outcomes. Third, a significant portion of the included studies lack sufficient statistical power to draw conclusive results to the broader populations. Finally, the lack of a standard definition of pharmaceutical outcomes, differences in participants characteristics, and the use of multiple medications across the different studies makes it difficult to compare findings and to perform meta-analysis.

Where associations between PGx-scores and treatment outcomes were established, effect size estimates (beta, odds ratios, hazard ratios) and measures of explained variance (R^2^) varied widely in studies included in our systematic review. For instance, the R^2^ of PGx-score models for predicting resistance to clozapine treatment with PGSSCZ in TRS individuals ranged from 2.03% ^36^ to 5.62% ^31^. Similarly, the reported odds ratios for clozapine response ranged from 1.94 [95%CI: 1.33 – 2.81] ^38^ to 6.50 [95%CI: 1.47 – 28.80] ^35^. These inconsistent findings can partly be explained by phenotypic heterogeneity, evident in diverse definitions and measurement of treatment outcomes and by differences in the sample size of these studies. As an example, the definition of TRS and TRD varies widely across studies ^36, 44, 56, 89–92^. Achieving uniformity in phenotype characterization, and harmonizing assessments across studies would help to improve the reliability of PGx-score for treatment outcomes.

Variation in sample size can also affect the size of individual study effect estimates and their statistical significance. Studies with small target or discovery samples, have limited statistical power to detect significant associations. Choi et al have demonstrated that in a discovery cohort of 100,000 samples, 200 to 500 samples in the target cohort are requisite to achieve 80% power for predicting traits across a spectrum of heritability estimates (*h*^2^:0.11 – 0.23) in polygenic models ^93^. Smaller sample sizes lead to larger sampling variance on individual marker effects and error accumulates across multiple markers such that the sample of variation on polygenic scores can be considerable. Recruiting a sufficiently large and well-characterized sample of uniformly treated individuals is a common challenge in PGx-score studies ^69, 94^.

### Future directions in PGx-score research

While PGx-scores hold promise for predicting treatment outcomes, they currently account for only a small proportion of the variance in treatment outcomes. This systematic review highlights the lack of well-defined phenotypes and small samples sizes that limit our ability to adequately quantify the genetic complexity associated with medication response. In this context, the following future directions may improve the predictive capacity of PGx-score and move us closer to their clinical utilization in psychiatry.

### Biologically informed PGx-scores

Previous PGx-score studies have been developed based on conventional polygenic modelling approaches, where the effect of genetic variants across the entire genome are aggregated, without taking into account the biological significance of these variants on the phenotype of interest ^72, 95^. A biology-informed polygenic score (B-PGS) model was introduced very recently as a novel approach to improve both the predictive capability and biological meaning of polygenic scores, while also reducing sequencing costs ^96, 97^. For example, in a study to predict psychosis, a pathway specific PGS that was restricted to genomic locations within “nervous system development” and “regulation of neuron differentiation”, explained a variance of 6.9% in the risk of psychosis, outperforming the conventional PGS where genome-wide SCZ variants accounted for only 3.7% ^98^. B-PGS potentially increase the polygenic signal to noise ratio by excluding variants with little association to pharmacogenomic outcomes and also enhance the clinical interpretability of polygenic models by focussing on specific molecular pathways ^99^. There is emerging evidence elsewhere in medicine that B-PGS may be useful for identification of new drug targets, for instance in inflammatory bowel disease ^100^.

### Multi-trait PGx-score

By leveraging the genetic correlation between multiple phenotypes, the multi-trait PGS approach aggregates genetic information across traits with the aim to improve the prediction power of PGx-scores ^101–103^. For example, in patients with BD, the polygenic scores of SCZ or MDD explained 0.80% ^47^ and 0.91% ^59^ of the variance in lithium response, respectively. Interestingly, combining the polygenic scores of SCZ and MDD, resulted in a better model, with an explained variance of 1.85% in lithium treatment response ^41^, indicating that multi-trait PGS outperform single trait PGS.

### Combining multimodal data and machine learning optimization

Researchers have begun to combine PGS with other data modalities, for example clinical and imaging data to improve model accuracy ^40, 104^. Machine learning methods are progressively being adopted for the analysis of multimodal or complex data comprising PGx-scores, socio-demographic, behavioural and clinical data ^105, 106^. This approach, exemplified in a few studies included in our review ^34, 40^, holds promising results for clinical translation. Nevertheless, replication of these complex studies is lacking and interpretation of machine learning algorithms could be difficult for clinicians, potentially limiting their acceptance ^107, 108^. To overcome this barrier, it is important for data scientists and clinicians to collaborate at an early stage of model development to ensure that these models are not only clinically useful but also calibrated and valid for local conditions and easily understandable for end users ^109–111^.

### Validation of polygenic models

Given the complexity of pharmacogenetic models, current sampling issues and the associated risks of false discovery and poor generalizability across different populations, external replication and validation of these models is critical for future implementation ^25, 112–114^. Only 26.4% of studies included in this systematic review employed external validation ^31, 33, 34, 36, 37, 43, 45, 46, 55, 56, 59, 62, 64, 115^.

### Multi-ancestry PGx-score

Nearly 90% of samples in the target and discovery cohorts of studies included in our systematic review were European descent. Genetic variations and their effect on treatment outcomes can vary significantly among different populations. Given the complex pattern of linkage disequilibrium (short genetic regions) and the significant difference in the frequency of genetic variants between populations, PGx-score constructed from one ancestral cohort may have lower prediction in another cohort ^112, 116, 117^. For instance in cardiovascular medicine, a Brazilian specific warfarin PGx-score used in a warfarin dosing algorithm was more accurate in Brazil, than the one developed in European population ^118^.

Conversely, polygenic models that incorporate information from ancestrally diverse populations, improve prediction performance particularly in underrepresented non-European populations ^119–123^. Diverse sampling is required to develop and validate more generalizable and transferable PGx-scores across diverse populations^72, 117^. These limitations hamper the translation of research findings into clinical practice and raise health disparity concerns. Thus, improving diversity in pharmacogenomic research is essential steps in creating polygenic models with broader application.

### Clinical implications of PGx-score

While it is clear that further development is required to improve accuracy of PGx-score and alone they have low clinical utility, findings are advancing our knowledge of pharmacogenomics toward better personalisation of treatment For instance, the genetic loading for SCZ demonstrates some capability to stratify individuals based on lithium treatment response in BD ^40, 41, 47^ and clozapine dosage in individuals with TRS ^33^.

Drawing parallels from other disciplines, such as cardiovascular medicine, PGS for coronary artery disease been used to reclassify patients from intermediate into high-risk categories translating into stronger statin use recommendations ^124, 125^. Similarly, genome-wide PGS in cardiovascular research have identified individuals with a four-fold increased risk, prompting recommendations for aggressive cholesterol-lowering therapy ^126^. Such evidence indicates that the polygenic scores have the potential to stratify patients, predict treatment outcomes and informed therapeutics decision making based on the genetic variation of population variation among different ancestral populations (Figure 4).

**Figure 4:**
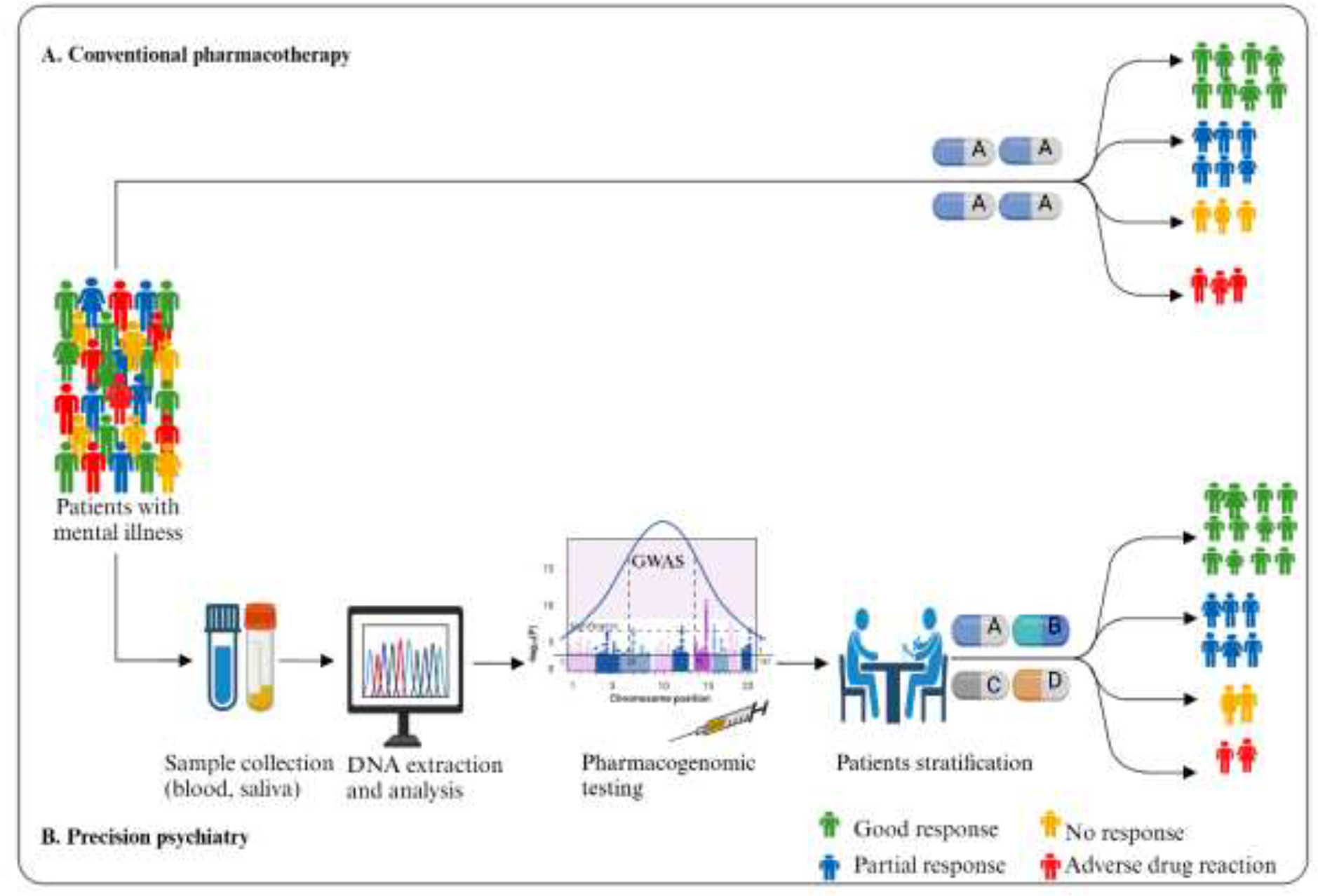
The potential use of PGx-score in precision psychiatry *Abbreviations:* DNA = Deoxyribonucleic acid

## Conclusions

In summary, this systematic review highlights that larger and more diverse target sample sizes, focussed on well-defined and standardised pharmacogenomic outcomes, with robust replication are required to optimise the development of PGx-scores. Currently the variance explained by these models is too small for effective clinical translation. However, new techniques, such as B-PGS and the use of multivariate modelling combining multiple traits PGS with clinical data look promising to the increase accuracy. Large scale consortia focused on pharmacogenomics are required to improve sample size and diversity.

## Data Availability

All data produced in the present work are contained in the manuscript

## References

1. Cuijpers P, Javed A, Bhui K. The WHO World Mental Health Report: a call for action. The British Journal of Psychiatry 2023; 222(6): 227–229.

2. GBD Mental Disorders Collaborators. Global, regional, and national burden of 12 mental disorders in 204 countries and territories, 1990–2019: a systematic analysis for the Global Burden of Disease Study 2019. The Lancet Psychiatry 2022; 9(2): 137-150.

3. Smith K, De Torres I. Mental health: A world of depression. Nature 2014; 515(181): 10–1038.

4. WHO. Nearly one billion people have a mental disorders 2022.

5. Patel V, Saxena S, Lund C, Thornicroft G, Baingana F, Bolton P et al. The Lancet Commission on global mental health and sustainable development. The lancet 2018; 392(10157): 1553–1598.

6. Health TLG. Mental health matters. The Lancet Global Health 2020; 8(11): e1352.

7. Nathan PE, Gorman JM. A guide to treatments that work. Oxford University Press 2015.

8. Hirschfeld RM. Efficacy of SSRIs and newer antidepressants in severe depression: comparison with TCAs. J Clin Psychiatry 1999; 60(5): 326–335.

9. Cipriani A, Furukawa TA, Salanti G, Chaimani A, Atkinson LZ, Ogawa Y et al. Comparative efficacy and acceptability of 21 antidepressant drugs for the acute treatment of adults with major depressive disorder: a systematic review and network meta-analysis. Focus 2018; 16(4): 420–429.

10. Hou L, Heilbronner U, Degenhardt F, Adli M, Akiyama K, Akula N et al. Genetic variants associated with response to lithium treatment in bipolar disorder: a genome-wide association study. The Lancet 2016; 387(10023): 1085–1093.

11. Siskind D, Orr S, Sinha S, Yu O, Brijball B, Warren N et al. Rates of treatment-resistant schizophrenia from first-episode cohorts: systematic review and meta-analysis. The British Journal of Psychiatry 2022; 220(3): 115–120.

12. McMahon FJ. Prediction of treatment outcomes in psychiatry--where do we stand ? Dialogues Clin Neurosci 2014; 16(4): 455–464.

13. Tansey KE, Guipponi M, Hu X, Domenici E, Lewis G, Malafosse A et al. Contribution of common genetic variants to antidepressant response. Biol Psychiatry 2013; 73(7): 679–682.

14. Crisafulli C, Fabbri C, Porcelli S, Drago A, Spina E, De Ronchi D et al. Pharmacogenetics of antidepressants. Front Pharmacol 2011; 2: 6.

15. Pardiñas AF, Owen MJ, Walters JTR. Pharmacogenomics: A road ahead for precision medicine in psychiatry. Neuron 2021; 109(24): 3914–3929.

16. Kang HJ, Kim KT, Yoo KH, Park Y, Kim JW, Kim SW et al. Genetic Markers for Later Remission in Response to Early Improvement of Antidepressants. Int J Mol Sci 2020; 21(14).

17. Lally J, Gaughran F, Timms P, Curran SR. Treatment-resistant schizophrenia: current insights on the pharmacogenomics of antipsychotics. Pharmgenomics Pers Med 2016; 9: 117–129.

18. Jeiziner C, Wernli U, Suter K, Hersberger KE, Meyer zu Schwabedissen HE. HLA-associated adverse drug reactions-scoping review. Clinical and Translational Science 2021; 14(5): 1648–1658.

19. Bousman CA, Jaksa P, Pantelis C. Systematic evaluation of commercial pharmacogenetic testing in psychiatry: a focus on CYP2D6 and CYP2C19 allele coverage and results reporting. Pharmacogenet Genomics 2017; 27(11): 387–393.

20. Licinio J, Wong ML. Pharmacogenomics of antidepressant treatment effects. Dialogues Clin Neurosci 2011; 13(1): 63–71.

21. Fagerness J, Fonseca E, Hess GP, Scott R, Gardner KR, Koffler M et al. Pharmacogenetic-guided psychiatric intervention associated with increased adherence and cost savings. The American journal of managed care 2014; 20(5): e146–156.

22. Relling MV, Dervieux T. Pharmacogenetics and cancer therapy. Nature reviews cancer 2001; 1(2): 99–108.

23. Lauschke VM, Ingelman-Sundberg M. Emerging strategies to bridge the gap between pharmacogenomic research and its clinical implementation. npj Genom Med 2020; 5.

24. Crouch DJ, Bodmer WF. Polygenic inheritance, GWAS, polygenic risk scores, and the search for functional variants. Proceedings of the National Academy of Sciences 2020; 117(32): 18924–18933.

25. Lee JW, Aminkeng F, Bhavsar A, Shaw K, Carleton B, Hayden M et al. The emerging era of pharmacogenomics: current successes, future potential, and challenges. Clinical genetics 2014; 86(1): 21–28.

26. Page MJ, McKenzie JE, Bossuyt PM, Boutron I, Hoffmann TC, Mulrow CD et al. The PRISMA 2020 statement: an updated guideline for reporting systematic reviews. International journal of surgery 2021; **88**: 105906.

27. Van Vugt L, Van den Reek J, Coenen M, de Jong E. A systematic review of pharmacogenetic studies on the response to biologics in patients with psoriasis. British Journal of Dermatology 2018; 178(1): 86–94.

28. Meerman JJ, Ter Hark SE, Janzing JGE, Coenen MJH. The Potential of Polygenic Risk Scores to Predict Antidepressant Treatment Response in Major Depression: A Systematic Review. J Affect Disord 2022; 304: 1–11.

29. Mayen-Lobo YG, Martinez-Magana JJ, Perez-Aldana BE, Ortega-Vazquez A, Genis-Mendoza AD, De Montellano D et al. Integrative Genomic-Epigenomic Analysis of Clozapine-Treated Patients with Refractory Psychosis. Pharmaceuticals 2021; 14(2): 16.

30. Campos AI, Ngo TT, Medland SE, Wray NR, Hickie IB, Byrne EM et al. Genetic risk for chronic pain is associated with lower antidepressant effectiveness: Converging evidence for a depression subtype. Aust N Z J Psychiatry 2022; 56(9): 1177–1186.

31. O’Connell KS, Koch E, Lenk HC, Akkouh IA, Hindley G, Jaholkowski P et al. Polygenic overlap with body-mass index improves prediction of treatment-resistant schizophrenia. Psychiatry Res 2023; 325: 115217.

32. Lin BD, Pinzon-Espinosa J, Blouzard E, van der Horst MZ, Okhuijsen-Pfeifer C, van Eijk KR et al. Associations Between Polygenic Risk Score Loading, Psychosis Liability, and Clozapine Use Among Individuals With Schizophrenia. JAMA PSYCHIATRY 2023; 80(2): 181–185.

33. Kappel DB, Legge SE, Hubbard L, Willcocks IR, O’Connell KS, Smith RL et al. Genomic Stratification of Clozapine Prescription Patterns Using Schizophrenia Polygenic Scores. Biol Psychiatry 2023; 93(2): 149–156.

34. Guo L-K, Su Y, Zhang Y-Y-N, Yu H, Lu Z, Li W-Q et al. Prediction of treatment response to antipsychotic drugs for precision medicine approach to schizophrenia: randomized trials and multiomics analysis. Military Medical Research 2023; 10(1): 24.

35. Talarico F, Costa GO, Ota VK, Santoro ML, Noto C, Gadelha A et al. Systems-Level Analysis of Genetic Variants Reveals Functional and Spatiotemporal Context in Treatment-resistant Schizophrenia. Mol Neurobiol 2022; 59(5): 3170–3182.

36. Pardinas AF, Smart SE, Willcocks IR, Holmans PA, Dennison CA, Lynham AJ et al. Interaction Testing and Polygenic Risk Scoring to Estimate the Association of Common Genetic Variants With Treatment Resistance in Schizophrenia. JAMA Psychiatry 2022; 79(3): 260–269.

37. Pain O, Hodgson K, Trubetskoy V, Ripke S, Marshe VS, Adams MJ et al. Identifying the Common Genetic Basis of Antidepressant Response. Biol Psychiatry Glob Open Sci 2022; 2(2): 115–126.

38. Okhuijsen-Pfeifer C, van der Horst MZ, Bousman CA, Lin B, van Eijk KR, Ripke S et al. Genome-wide association analyses of symptom severity among clozapine-treated patients with schizophrenia spectrum disorders. Transl Psychiatr 2022; 12(1): 145.

39. Facal F, Arrojo M, Paz E, Paramo M, Costas J. Association between psychiatric hospitalizations of patients with schizophrenia and polygenic risk scores based on genes with altered expression by antipsychotics. Acta Psychiatr Scand 2022; 146(2): 139–150.

40. Cearns M, Amare AT, Schubert KO, Thalamuthu A, Frank J, Streit F et al. Using polygenic scores and clinical data for bipolar disorder patient stratification and lithium response prediction: machine learning approach - CORRIGENDUM. Br J Psychiatry 2022; 221(2): 494.

41. Schubert KO, Thalamuthu A, Amare AT, Frank J, Streit F, Adl M, et al. Combining schizophrenia and depression polygenic risk scores improves the genetic prediction of lithium response in bipolar disorder patients. Transl Psychiatr 2021; 11(1): 606.

42. Werner MCF, Wirgenes KV, Haram M, Bettella F, Lunding SH, Rodevand L et al. Indicated association between polygenic risk score and treatment-resistance in a naturalistic sample of patients with schizophrenia spectrum disorders. Schizophr Res 2020; 218: 55–62.

43. Zhang JP, Robinson D, Yu J, Gallego J, Fleischhacker WW, Kahn RS, et al. Schizophrenia Polygenic Risk Score as a Predictor of Antipsychotic Efficacy in First-Episode Psychosis. Am J Psychiat 2019; 176(1): 21-28.

44. Gasse C, Wimberley T, Wang Y, Mors O, Borglum A, Als TD et al. Schizophrenia polygenic risk scores, urbanicity and treatment-resistant schizophrenia. Schizophr Res 2019; 212: 79–85.

45. Santoro ML, Ota V, de Jong S, Noto C, Spindola LM, Talarico F, et al. Polygenic risk score analyses of symptoms and treatment response in an antipsychotic-naive first episode of psychosis cohort. Transl Psychiatr 2018; 8(1): 174.

46. Li J, Yoshikawa A, Brennan MD, Ramsey TL, Meltzer HY. Genetic predictors of antipsychotic response to lurasidone identified in a genome wide association study and by schizophrenia risk genes. Schizophr Res 2018; 192: 194–204.

47. International Consortium on Lithium G, Amare AT, Schubert KO, Hou L, Clark SR, Papiol S et al. Association of Polygenic Score for Schizophrenia and HLA Antigen and Inflammation Genes With Response to Lithium in Bipolar Affective Disorder: A Genome-Wide Association Study. JAMA Psychiatry 2018; 75(1): 65-74.

48. Yoshida K, Marshe VS, Elsheikh SSM, Maciukiewicz M, Tiwari AK, Brandl EJ et al. Polygenic risk scores analyses of psychiatric and metabolic traits with antipsychotic- induced weight gain in schizophrenia: an exploratory study. Pharmacogenomics J 2023.

49. Muntane G, Vazquez-Bourgon J, Sada E, Martorell L, Papiol S, Bosch E et al. Polygenic risk scores enhance prediction of body mass index increase in individuals with a first episode of psychosis. Eur Psychiatry 2023; 66(1): e28.

50. Lacaze P, Ronaldson KJ, Zhang EJ, Alfirevic A, Shah H, Newman L et al. Genetic associations with clozapine-induced myocarditis in patients with schizophrenia. Transl Psychiatr 2020; 10(1): 37.

51. Blackman RK, Dickinson D, Eisenberg DP, Gregory MD, Apud JA, Berman KF. Antipsychotic medication-mediated cognitive change in schizophrenia and polygenic score for cognitive ability. Schizophr Res-Cogn 2022; 27: 7.

52. Segura À G, Martínez-Pinteño A, Gassó P, Rodríguez N, Bioque M, Cuesta MJ et al. Metabolic polygenic risk scores effect on antipsychotic-induced metabolic dysregulation: A longitudinal study in a first episode psychosis cohort. Schizophr Res 2022; 244: 101–110.

53. Gendep Investigators, Investigators M, Investigators SD. Common genetic variation and antidepressant efficacy in major depressive disorder: a meta-analysis of three genome-wide pharmacogenetic studies. American Journal of Psychiatry 2013; 170(2): 207–217.

54. Amare AT, Schubert KO, Tekola-Ayele F, Hsu YH, Sangkuhl K, Jenkins G et al. Association of the Polygenic Scores for Personality Traits and Response to Selective Serotonin Reuptake Inhibitors in Patients with Major Depressive Disorder. Front Psychiatry 2018; 9: 65.

55. Marshe VS, Maciukiewicz M, Hauschild AC, Islam F, Qin L, Tiwari AK et al. Genome-wide analysis suggests the importance of vascular processes and neuroinflammation in late-life antidepressant response. Transl Psychiatr 2021; 11(1): 127.

56. Amare AT, Schubert KO, Tekola-Ayele F, Hsu YH, Sangkuhl K, Jenkins G et al. The association of obesity and coronary artery disease genes with response to SSRIs treatment in major depression. Journal of Neural Transmission 2019; 126(**1**): 35–45.

57. Men X, Marshe V, Elsheikh SS, Alexopoulos GS, Marino P, Meyers BS et al. Genomic Investigation of Remission and Relapse of Psychotic Depression Treated with Sertraline plus Olanzapine: The STOP-PD II Study. Neuropsychobiology 2023; 82(3): 168–178.

58. Zwicker A, Fabbri C, Rietschel M, Hauser J, Mors O, Maier W et al. Genetic disposition to inflammation and response to antidepressants in major depressive disorder. Journal of Psychiatric Research 2018; 105: 17–22.

59. Amare AT, Schubert KO, Hou L, Clark SR, Papiol S, Cearns M, et al. Association of polygenic score for major depression with response to lithium in patients with bipolar disorder. Mol Psychiatr 2021; 26(6): 2457-2470.

60. Coombes BJ, Millischer V, Batzler A, Larrabee B, Hou L, Papiol S, et al. Association of Attention-Deficit/Hyperactivity Disorder and Depression Polygenic Scores with Lithium Response: A Consortium for Lithium Genetics Study. Complex Psychiatry 2021; 7(3-4): 80-89.

61. Millischer V, Matheson GJ, Bergen SE, Coombes BJ, Ponzer K, Wikström F et al. Improving lithium dose prediction using population pharmacokinetics and pharmacogenomics: a cohort genome-wide association study in Sweden. The Lancet Psychiatry 2022; 9(6): 447–457.

62. Meijs H, Prentice A, Lin BD, De Wilde B, Van Hecke J, Niemegeers P et al. A polygenic-informed approach to a predictive EEG signature empowers antidepressant treatment prediction: A proof-of-concept study. Eur Neuropsychopharmacol 2022; 62: 49–60.

63. Guo W, Machado-Vieira R, Mathew S, Murrough JW, Charney DS, Gruenbaum M et al. Exploratory genome-wide association analysis of response to ketamine and a polygenic analysis of response to scopolamine in depression. Translational Psychiatry 2018; 8: 9.

64. Amare AT, Thalamuthu A, Schubert KO, Fullerton JM, Ahmed M, Hartmann S et al. Association of polygenic score and the involvement of cholinergic and glutamatergic pathways with lithium treatment response in patients with bipolar disorder. Mol Psychiatr 2023.

65. Cardno AG, Gottesman II. Twin studies of schizophrenia: from bow-and-arrow concordances to star wars Mx and functional genomics. American journal of medical genetics 2000; 97(1): 12–17.

66. Yang J, Zeng J, Goddard ME, Wray NR, Visscher PM. Concepts, estimation and interpretation of SNP-based heritability. Nature genetics 2017; 49(9): 1304–1310.

67. Trubetskoy V, Pardiñas AF, Qi T, Panagiotaropoulou G, Awasthi S, Bigdeli TB et al. Mapping genomic loci implicates genes and synaptic biology in schizophrenia. Nature 2022; 604(7906): 502-508.

68. Ripke S, Neale BM, Corvin A, Walters JTR, Farh K-H, Holmans PA et al. Biological insights from 108 schizophrenia-associated genetic loci. Nature 2014; 511(7510): 421-427.

69. Dudbridge F. Power and predictive accuracy of polygenic risk scores. PLoS genetics 2013; 9(3): e1003348.

70. Purcell SM, Wray NR, Stone JL, Visscher PM, O’Donovan MC, Sullivan PF et al. Common polygenic variation contributes to risk of schizophrenia and bipolar disorder. Nature 2009; 460(7256): 748-752.

71. Pantelis C, Papadimitriou GN, Papiol S, Parkhomenko E, Pato MT, Paunio T et al. Biological insights from 108 schizophrenia-associated genetic loci. Nature 2014; 511(7510): 421-427.

72. Duncan L, Shen H, Gelaye B, Meijsen J, Ressler K, Feldman M et al. Analysis of polygenic risk score usage and performance in diverse human populations. Nature Communications 2019; 10(1): 3328.

73. Sekar A, Bialas AR, de Rivera H, Davis A, Hammond TR, Kamitaki N et al. Schizophrenia risk from complex variation of complement component 4. Nature 2016; 530(7589): 177-183.

74. Satterthwaite TD, Wolf DH, Calkins ME, Vandekar SN, Erus G, Ruparel K et al. Structural Brain Abnormalities in Youth With Psychosis Spectrum Symptoms. JAMA Psychiatry 2016; 73(5): 515–524.

75. Vissink CE, Winter-van Rossum I, Cannon TD, Fusar-Poli P, Kahn RS, Bossong MG. Structural Brain Volumes of Individuals at Clinical High Risk for Psychosis: A Meta-analysis. Biological Psychiatry Global Open Science 2022; 2(2): 147–152.

76. Mouchlianitis E, McCutcheon R, Howes OD. Brain-imaging studies of treatment-resistant schizophrenia: a systematic review. Lancet Psychiatry 2016; 3(5): 451–463.

77. Hagenaars SP, Coleman JRI, Choi SW, Gaspar H, Adams MJ, Howard DM et al. Genetic comorbidity between major depression and cardio-metabolic traits, stratified by age at onset of major depression. Am J Med Genet B Neuropsychiatr Genet 2020; 183(6): 309–330.

78. Amare AT, Schubert KO, Klingler-Hoffmann M, Cohen-Woods S, Baune BT. The genetic overlap between mood disorders and cardiometabolic diseases: a systematic review of genome wide and candidate gene studies. Translational psychiatry 2017; 7(1): e1007–e1007.

79. Willer C, Speliotes E, Loos R, Li S, Lindgren C, Heid I et al. Genetic Investigation of ANthropometric Traits Consortium Six new loci associated with body mass index highlight a neuronal influence on body weight regulation. Nat Genet 2009; 41(1): 25.

80. Maccarrone G, Ditzen C, Yassouridis A, Rewerts C, Uhr M, Uhlen M et al. Psychiatric patient stratification using biosignatures based on cerebrospinal fluid protein expression clusters. Journal of psychiatric research 2013; 47(11): 1572–1580.

81. Okbay A, Baselmans BML, De Neve J-E, Turley P, Nivard MG, Fontana MA et al. Genetic variants associated with subjective well-being, depressive symptoms, and neuroticism identified through genome-wide analyses. Nature Genetics 2016; 48(6): 624–633.

82. Lo MT, Hinds DA, Tung JY, Franz C, Fan CC, Wang Y et al. Genome-wide analyses for personality traits identify six genomic loci and show correlations with psychiatric disorders. Nat Genet 2017; 49(1): 152–156.

83. Hayward M, Moran P. Comorbidity of personality disorders and mental illnesses. Psychiatry 2008; 7(3): 102–104.

84. Kohli R. A Systematic Review To Evaluate The Association Between Medication Adherence And Personality Traits. Value in Health 2017; 20(9): A686.

85. Schulze TG, Alda M, Adli M, Akula N, Ardau R, Bui ET et al. The International Consortium on Lithium Genetics (ConLiGen): an initiative by the NIMH and IGSLI to study the genetic basis of response to lithium treatment. Neuropsychobiology 2010; 62(1): 72–78.

86. Pardiñas AF, Kappel DB, Roberts M, Tipple F, Shitomi-Jones LM, King A et al. Pharmacokinetics and pharmacogenomics of clozapine in an ancestrally diverse sample: a longitudinal analysis and genome-wide association study using UK clinical monitoring data. The Lancet Psychiatry 2023; 10(3): 209–219.

87. Cearns M, Opel N, Clark S, Kaehler C, Thalamuthu A, Heindel W, et al. Predicting rehospitalization within 2 years of initial patient admission for a major depressive episode: a multimodal machine learning approach. Transl Psychiatr 2019; 9(1): 285.

88. Dziopa K, Chaturvedi N, Vugt M, Gratton J, Maclean R, Hingorani A, et al. Combining stacked polygenic scores with clinical risk factors improves cardiovascular risk prediction in people with type 2 diabetes. medRxiv 2022: 2022.2009. 2001.22279477.

89. Fanelli G, Domschke K, Minelli A, Gennarelli M, Martini P, Bortolomasi M et al. A meta-analysis of polygenic risk scores for mood disorders, neuroticism, and schizophrenia in antidepressant response. Eur Neuropsychopharmacol 2022; 55: 86–95.

90. Taylor RW, Coleman JRI, Lawrence AJ, Strawbridge R, Zahn R, Cleare AJ. Predicting clinical outcome to specialist multimodal inpatient treatment in patients with treatment resistant depression. J Affect Disord 2021; 291: 188–197.

91. Fanelli G, Benedetti F, Kasper S, Zohar J, Souery D, Montgomery S et al. Higher polygenic risk scores for schizophrenia may be suggestive of treatment non-response in major depressive disorder. Prog Neuropsychopharmacol Biol Psychiatry 2021; 108: 110170.

92. Wigmore EM, Hafferty JD, Hall LS, Howard DM, Clarke TK, Fabbri C et al. Genome-wide association study of antidepressant treatment resistance in a population-based cohort using health service prescription data and meta-analysis with GENDEP. Pharmacogenomics J 2020; 20(2): 329–341.

93. Choi SW, Mak TS, O’Reilly PF. Tutorial: a guide to performing polygenic risk score analyses. Nat Protoc 2020; 15(9): 2759–2772.

94. Blagec K, Swen JJ, Koopmann R, Cheung K-C, Crommentuijn-van Rhenen M, Holsappel I et al. Pharmacogenomics decision support in the U-PGx project: Results and advice from clinical implementation across seven European countries. PloS one 2022; 17(6): e0268534.

95. Wand H, Lambert SA, Tamburro C, Iacocca MA, O’Sullivan JW, Sillari C et al. Improving reporting standards for polygenic scores in risk prediction studies. Nature 2021; 591(7849): 211-219.

96. Hari Dass SA, McCracken K, Pokhvisneva I, Chen LM, Garg E, Nguyen TTT et al. A biologically-informed polygenic score identifies endophenotypes and clinical conditions associated with the insulin receptor function on specific brain regions. EBioMedicine 2019; 42: 188–202.

97. Scharfe CPI, Tremmel R, Schwab M, Kohlbacher O, Marks DS. Genetic variation in human drug-related genes. Genome Med 2017; 9(1): 117.

98. Tubbs JD, Leung PBM, Zhong Y, Zhan N, Hui TCK, Ho KKY et al. Pathway-Specific Polygenic Scores Improve Cross-Ancestry Prediction of Psychosis and Clinical Outcomes. medRxiv 2023: 2023.2009. 2001.23294957.

99. Bennett D, O’Shea D, Ferguson J, Morris D, Seoighe C. Controlling for background genetic effects using polygenic scores improves the power of genome-wide association studies. Scientific Reports 2021; 11(1): 19571.

100. Bodea CA, Macoritto M, Liu Y, Zhang W, Karman J, King EA et al. Pathway specific polygenic risk scores identify pathways and patient clusters associated with inflammatory bowel disease risk, severity and treatment response. medRxiv 2021: 2021.2011. 2019.21266549.

101. Krapohl E, Patel H, Newhouse S, Curtis CJ, von Stumm S, Dale PS et al. Multi-polygenic score approach to trait prediction. Mol Psychiatr 2018; 23(5): 1368–1374.

102. Maier RM, Zhu Z, Lee SH, Trzaskowski M, Ruderfer DM, Stahl EA et al. Improving genetic prediction by leveraging genetic correlations among human diseases and traits. Nature communications 2018; 9(1): 989.

103. Albiñana C, Zhu Z, Schork AJ, Ingason A, Aschard H, Brikell I et al. Multi-PGS enhances polygenic prediction by combining 937 polygenic scores. Nature Communications 2023; 14(1): 4702.

104. Wang M, Hu K, Fan L, Yan H, Li P, Jiang T et al. Predicting treatment response in schizophrenia with magnetic resonance imaging and polygenic risk score. Front Genet 2022; 13: 848205.

105. Zou J, Huss M, Abid A, Mohammadi P, Torkamani A, Telenti A. A primer on deep learning in genomics. Nature genetics 2019; 51(1): 12–18.

106. Eraslan G, Avsec Ž, Gagneur J, Theis FJ. Deep learning: new computational modelling techniques for genomics. Nature Reviews Genetics 2019; 20(7): 389–403.

107. Wagner MW, Namdar K, Biswas A, Monah S, Khalvati F, Ertl-Wagner BB. Radiomics, machine learning, and artificial intelligence—what the neuroradiologist needs to know. Neuroradiology 2021: 1–11.

108. Ngiam KY, Khor W. Big data and machine learning algorithms for health-care delivery. The Lancet Oncology 2019; 20(5): e262–e273.

109. Cearns M, Hahn T, Baune BT. Recommendations and future directions for supervised machine learning in psychiatry. Translational Psychiatry 2019; 9(1): 271.

110. Winter NR, Cearns M, Clark SR, Leenings R, Dannlowski U, Baune BT et al. From multivariate methods to an AI ecosystem. Molecular Psychiatry 2021; 26(11): 6116–6120.

111. Jarrett D, Stride E, Vallis K, Gooding MJ. Applications and limitations of machine learning in radiation oncology. The British journal of radiology 2019; 92(1100): 20190001.

112. Mostafavi H, Harpak A, Agarwal I, Conley D, Pritchard JK, Przeworski M. Variable prediction accuracy of polygenic scores within an ancestry group. elife 2020; 9: e48376.

113. Kerminen S, Martin AR, Koskela J, Ruotsalainen SE, Havulinna AS, Surakka I et al. Geographic variation and bias in the polygenic scores of complex diseases and traits in Finland. The American Journal of Human Genetics 2019; 104(6): 1169–1181.

114. Moons KG, Kengne AP, Woodward M, Royston P, Vergouwe Y, Altman DG et al. Risk prediction models: I. Development, internal validation, and assessing the incremental value of a new (bio) marker. Heart 2012; 98(9): 683–690.

115. Maciukiewicz M, Tiwari AK, Zai CC, Gorbovskaya I, Laughlin CP, Nurmi EL et al. Genome-wide association study on antipsychotic-induced weight gain in Europeans and African-Americans. Schizophrenia Research 2019; 212: 204–212.

116. Kuchenbaecker K, Telkar N, Reiker T, Walters RG, Lin K, Eriksson A et al. The transferability of lipid loci across African, Asian and European cohorts. Nature communications 2019; 10(1): 4330.

117. Martin AR, Kanai M, Kamatani Y, Okada Y, Neale BM, Daly MJ. Clinical use of current polygenic risk scores may exacerbate health disparities. Nature Genetics 2019; 51(4): 584–591.

118. Perini J, Struchiner C, Silva-Assunção E, Santana I, Rangel F, Ojopi E et al. Pharmacogenetics of warfarin: development of a dosing algorithm for Brazilian patients. Clinical Pharmacology & Therapeutics 2008; 84(6): 722–728.

119. Weissbrod O, Kanai M, Shi H, Gazal S, Peyrot WJ, Khera AV, et al. Leveraging fine-mapping and non-European training data to improve cross-population polygenic risk scores. MedRxiv 2021: 2021.2001. 2019.21249483.

120. Márquez-Luna C, Loh PR, Consortium SATD, Consortium STD, Price AL. Multiethnic polygenic risk scores improve risk prediction in diverse populations. Genetic epidemiology 2017; 41(8): 811–823.

121. Weissbrod O, Kanai M, Shi H, Gazal S, Peyrot WJ, Khera AV et al. Leveraging fine-mapping and multipopulation training data to improve cross-population polygenic risk scores. Nature Genetics 2022; 54(4): 450–458.

122. Ruan Y, Lin Y-F, Feng Y-CA, Chen C-Y, Lam M, Guo Z et al. Improving polygenic prediction in ancestrally diverse populations. Nature genetics 2022; 54(5): 573–580.

123. Patel AP, Wang M, Ruan Y, Koyama S, Clarke SL, Yang X et al. A multi-ancestry polygenic risk score improves risk prediction for coronary artery disease. Nat Med 2023; 29(7): 1793–1803.

124. Kullo IJ, Jouni H, Austin EE, Brown S-A, Kruisselbrink TM, Isseh IN et al. Incorporating a genetic risk score into coronary heart disease risk estimates: effect on low-density lipoprotein cholesterol levels (the MI-GENES Clinical Trial). Circulation 2016; 133(12): 1181–1188.

125. Natarajan P, Young R, Stitziel NO, Padmanabhan S, Baber U, Mehran R et al. Polygenic risk score identifies subgroup with higher burden of atherosclerosis and greater relative benefit from statin therapy in the primary prevention setting. Circulation 2017; 135(22): 2091–2101.

126. Khera AV, Chaffin M, Aragam KG, Haas ME, Roselli C, Choi SH et al. Genome-wide polygenic scores for common diseases identify individuals with risk equivalent to monogenic mutations. Nat Genet 2018; 50(9): 1219–1224.

