## Supplementary file 1 for "Pharmacogenomic scores in psychiatry: Systematic review of existing evidence"

**Supplementary Material**

**Supplementary Table 1:** Complete search strategies for the association of pharmacogenomics polygenic scores and treatment outcomes in psychiatry practice

| Concepts | Search strings (PubMed) | PubMed | EMBASE | Web of Science |
| --- | --- | --- | --- | --- |
| Polygenic score (#1) | “Polygenic score*”[tiab] OR “Polygenic risk score*”[tiab] OR PRS OR “Risk profile score*”[tiab] OR “Genetic risk score*”[tiab] OR “Gene score*”[tiab] OR “Genetic score*”[tiab] OR polygenic*[tiab] OR "Pharmacogenomic Variants"[Mesh] OR "Pharmacogenomic Testing"[Mesh] OR Pharmaco-omic*[tiab] OR pharmacogeno*[tiab] OR "Pharmacogenetics"[Mesh] | 37,106 | 68,712 | 39,362 |
| Psychotropic drugs (#2) | "Antipsychotic Agents"[Mesh] OR "Antipsychotic Agents" [Pharmacological Action] OR antipsycho*[tiab] OR "Antidepressive Agents"[Mesh] Antidepress*[tiab] OR "Antidepressive Agents" [Pharmacological Action] OR "Anti-Anxiety Agents"[Mesh] OR Anti-Anxiet*[tiab] OR Antixiolytic*[tiab] OR Valproic acid[tiab] OR Valproate[tiab] OR Divalproate[tiab] OR Divalproex[tiab] OR Carbamazepine[tiab] OR Oxcarbazepine[tiab] OR Risperidone[tiab] OR Gabapentin[tiab] OR Lamotrigine[tiab] OR Licarbazepine[tiab] OR Pregabalin[tiab] OR Tiagabine[tiab] OR Zonisamide[tiab] OR Lithium[tiab] | 153,800 | 449,6049 | 385,309 |
|  | 1 # 2 | 1,005 | 1735 | 2149 |
|  | Studies included after-duplication | 3,303 | | |
|  | Studies included after title and abstracts | 127 | | |
|  | Records identified from citation searching | 6 | | |
|  | Studies included after full-text review | 53 | | |

**PubMed search string**

("Polygenic score*"[tiab] OR "Polygenic risk score*"[tiab] OR PRS OR "Risk profile score*"[tiab] OR "Genetic risk score*"[tiab] OR "Gene score*"[tiab] OR "Genetic score*"[tiab] OR polygenic*[tiab] OR "Pharmacogenomic Variants"[Mesh] OR "Pharmacogenomic Testing"[Mesh] OR Pharmaco-omic*[tiab] OR pharmacogeno*[tiab] OR "Pharmacogenetics"[Mesh] AND ((humans[Filter]) AND (english[Filter]))) AND ("Antipsychotic Agents"[Mesh] OR "Antipsychotic Agents" [Pharmacological Action] OR antipsycho*[tiab] OR "Antidepressive Agents"[Mesh] Antidepress*[tiab] OR "Antidepressive Agents" [Pharmacological Action] OR "Anti-Anxiety Agents"[Mesh] OR Anti-Anxiet*[tiab] OR Valproic acid[tiab] OR Valproate[tiab] OR Divalproate[tiab] OR Divalproex[tiab] OR Carbamazepine[tiab] OR Oxcarbazepine[tiab] OR Risperidone[tiab] OR Gabapentin[tiab] OR Lamotrigine[tiab] OR Licarbazepine[tiab] OR Pregabalin[tiab] OR Tiagabine[tiab] OR Zonisamide[tiab] OR Lithium[tiab] AND ((humans[Filter]) AND (english[Filter])))

**Embase (Ovid platform) search string**
(("Pharmacogenomic Variants" or "Pharmacogenomic Testing" or Pharmacogenetics or "Polygenic score*" or "Polygenic risk score*" or PRS or "Risk profile score*" or "Genetic risk score*" or "Gene score*" or "Genetic score*" or polygenic* or Pharmaco-omic* or pharmacogeno*) AND ("Antipsychotic Agents" or "Antidepressive Agents" or "Antidepressive Agents" or "Anti-Anxiety Agents" or antipsycho* or Antidepress* or Anti-Anxiet* or Valproic acid or Valproate or Divalproate or Divalproex or Carbamazepine or Oxcarbazepine or Risperidone or Gabapentin or Lamotrigine or Licarbazepine or Pregabalin or Tiagabine or Zonisamide or Lithium)).mp

**Web of Science**

TS=(("Pharmacogenomic Variants" OR "Pharmacogenomic Testing" OR Pharmacogenetics OR "Polygenic score*" OR "Polygenic risk score*" OR PRS OR "Risk profile score*" OR "Genetic risk score*" OR "Gene score*" OR "Genetic score*" OR polygenic* OR Pharmaco-omic* OR pharmacogene) AND ("Antipsychotic Agents" OR "Antidepressive Agents" OR "Antidepressive Agents" OR "Anti-Anxiety Agents" OR antipsycho* OR Antidepress* OR Anti-Anxiet* OR Valproic acid OR Valproate OR Divalproate OR Divalproex OR Carbamazepine OR Oxcarbazepine OR Risperidone OR Gabapentin OR Lamotrigine OR Licarbazepine OR Pregabalin OR Tiagabine OR Zonisamide OR Lithium))

**Supplementary Table 2:** Quality assessment of studies on the association between PGx-score and treatment outcomes

| Author (year) | Q1 | Q2 | Q3 | Q4 | Q5 | Q6 | Q7 | Q8 |
| --- | --- | --- | --- | --- | --- | --- | --- | --- |
| *Yoshida et al., (2023)* | Y | N | Y | N | Y | Y | Linear and logistic regression | Y |
| *O’Connell et al., (2023)* | Y | N | Y | Y | Y | Y | Chi-square | Y |
| *Muntane et al., (2023)* | Y | N | Y | N | Y | Y | Multiple linear regression and chi-square | Y |
| *Morgenroth et al., (2023)* | Y | N | Y | N | Y | Y | Linear and logistic regression | Y |
| *Lin et al., (2023)* | Y | N | Y | N | Y | Y | Multinomial logistic regression | Y |
| *Kappel et al., (2023)* | Y | Y | Y | Y | Y | Y | Binomial and multinomial logistic regression | Y |
| *Men et al., (2023)* | Y | N | Y | N | Y | Y | Logistic regression | Y |
| *Guo et al., (2023)* | Y | N | Y | Y | Y | Y | Machine learning analysis | Y |
| *Amare et al., (2023)* | Y | N | Y | Y | Y | Y | Linear and logistic regression | Y |
| *Talarico et al., (2022)* | Y | N | Y | N | N | Y | Logistic regression analysis | Y |
| *Segura et al., (2022)* | Y | N | Y | N | Y | Y | Linear mixed effects | Y |
| *Pardinas et al., (2022)* | Y | Y | Y | Y | Y | Y | Logistic regression analysis & Meta-analysis | Y |
| *Pain et al., (2022)* | Y | Y | Y | Y | Y | Y | Linear regression and meta-analysis | Y |
| *Okhuijsen-Pfeifer et al., (2022)* | N | N | Y | N | Y | Y | Logistic regression analysis | Y |
| *Nøhr et al., (2022)* | N | N | Y | N | Y | Y | Linear regression | Y |
| *Millischer et al., (2022)* | Y | N | Y | N | Y | Y | Linear mixed effect | Y |
| *Meijs et al., (2022)* | Y | N | Y | Y | Y | Y | Linear regression | Y |
| *Lu et al., (2022)* | N | N | Y | N | Y | Y | Linear and logistic regression | Y |
| *Fanelli et al., (2022)* | Y | Y | Y | N | Y | Y | Linear regression and meta-analysis | Y |
| *Facal et al., (2022)* | Y | N | Y | N | Y | Y | Logistic regression | Y |
| *Cearns et al., (2022)* | N | N | Y | N | Y | Y | Regularized linear (Ridge and Elastic-net and random forest analysis | Y |
| *Campos et al., (2022* | Y | N | Y | N | Y | Y | Logistic regression | Y |
| *Blackman et al., (2022)* | N | N | Y | N | Y | Y | Linear regression | N |
| *Taylor et al., (2021)* | Y | N | Y | N | N | Y | Elastic-net logistic regression | Y |
| *Schubert et al., (2021)* | Y | Y | Y | N | Y | Y | Linear and logistic regression | Y |
| *Mayen-Lobo et al., (2021)* | Y | N | Y | N | Y | Y | Logistic regression | Y |
| *Marshe et al., (2021)* | Y | N | Y | Y | Y | Y | Linear and logistic regression analysis | Y |
| *Kowalec et al., (2021)* | N | N | Y | N | N | N | Logistic regression | Y |
| *Hommers et al., (2021)* | N | N | Y | N | Y | N | Logistic regression | N |
| *Fanelli et al., (2021)* | Y | Y | Y | N | Y | Y | Regression analysis | Y |
| *Coombes et al., (2021)* | Y | N | Y | N | Y | Y | Linear and logistic regression | Y |
| *Amare et al., (2021)* | Y | N | Y | Y | Y | Y | Logistic and linear regression | Y |
| *Wigmore et al., (2020)* | Y | Y | Y | N | Y | Y | Linear mixed model analysis | Y |
| *Werner et al., (2020)* | N | N | Y | N | Y | Y | Logistic regression | Y |
| *Li et al., (2020)* | N | N | Y | N | Y | Y | Not specified | N |
| *Lacaze et al., (2020)* | N | N | Y | N | Y | Y | Logistic regression | Y |
| *Zhang et al., (2019)* | Y | N | Y | Y | Y | Y | Linear regression | Y |
| *Maciukiewicz et al., (2019)* | Y | N | Y | Y | Y | Y | Linear regression | Y |
| *Gasse et al., (2019)* | N | N | Y | N | Y | N | Hazard ratio | Y |
| *Amare et al., (2019)* | Y | N | Y | Y | Y | Y | Logistic regression analysis | Y |
| *Zwicker et al., (2018)* | Y | N | Y | N | Y | Y | Mixed effects model f | Y |
| *Ward et al., (2018)* | N | N | Y | N | N | Y | Random effects meta-analysis | N |
| *Santoro et al., 2018)* | Y | N | Y | Y | Y | Y | Linear regression analysis | Y |
| *Li et al., (2018)* | N | N | Y | Y | Y | Y | Logistic regression | Y |
| *Guo et al., (2018)* | Y | N | Y | N | N | Y | Linear regression analysis | Y |
| *Amare, et al., (2018)* | Y | N | Y | N | Y | Y | Logistic regression analysis | Y |
| *International Consortium on Lithium, G., et al., (2018)* | Y | N | Y | N | Y | Y | Linear and logistic regression and meta-analysis | Y |
| *Wimberley et al., (2017)* | Y | N | Y | N | Y | N | Logistic regression | Y |
| *Garcia-Gonzalez et al., (2017)* | Y | Y | Y | N | Y | Y | Linear regression analysis in each cohort, followed by a fixed effect meta-analysis | Y |
| *Martin & Mowry, (2016)* | N | N | Y | N | N | Y | Logistic regression analysis | Y |
| *Hettige et al., (2016)* | Y | N | Y | N | N | Y | Linear regression | Y |
| *Tansey et al., (2014)* | Y | Y | Y | N | N | Y | Linear regression analysis | Y |
| *Gendep Invesigators et al (2013)* | Y | Y | Y | N | N | Y | Linear and logistic regression | Y |

**Legends:**

**Y = Yes; N = No**

**Q1** = Was there a clear rationale for the selected PGS?

**Q2** = Was a power calculation performed?

**Q3** = Were the in- and exclusion criteria clearly described?

**Q4** = Was there an external validation cohort?

**Q5** = Was adjustment for multiple testing applied?

**Q6** = Are the methods used to construct the PGS specified?

**Q7** = What type of analysis was used for the main comparison?

**Q8** = Were confounders or covariates considered in the analyses?
